# Air pollution exposure in Generation Scotland: molecular fingerprints and health outcomes

**DOI:** 10.64898/2026.03.04.26347573

**Authors:** Josephine A. Robertson, Ilse Krätschmer, Anne Richmond, Daniel L. McCartney, Jakub Bajzik, Spyros Vernardis, Janie Corley, Samuel J. Tomlinson, Massimo Vieno, Aleksandra D. Chybowska, Arturas Grauslys, Hannah M. Smith, Charles Brigden, Christoph B. Messner, Aleksej Zelezniak, Markus Ralser, Tom C Russ, Jamie Pearce, Simon R. Cox, Matthew R. Robinson, Riccardo E. Marioni

**Affiliations:** Institute of Genetics and Cancer, University of Edinburgh, UK; Institute of Science and Technology, Klosterneuburg, Austria; The Francis Crick Institute, Molecular Biology of Metabolism Laboratory, London, UK; Eliptica Limited, The London Cancer Hub, Cotswold Road, Sutton, London, UK; The Lothian Birth Cohorts, Department of Psychology, University of Edinburgh, UK; UK Centre for Ecology and Hydrology, Lancaster Environment Centre, Lancaster, UK; UK Centre for Ecology and Hydrology, Edinburgh, UK; Precision Proteomics Center, Swiss Institute of Allergy and Asthma Research, University of Zurich, Switzerland; Randall Centre for Cell & Molecular Biophysics, King’s College London, New Hunt’s House, Guy’s Campus, SE1 1UL London, UK; Department of Biology and Biological Engineering, Chalmers University of Technology, Kemivägen 10, SE-412 96, Gothenburg, Sweden; Institute of Biotechnology, Life Sciences Centre, Vilnius University, Sauletekio al. 7, LT10257 Vilnius, Lithuania; Department of Biochemistry, Charité Universitätsmedizin Berlin, Berlin, Germany; Division of Psychiatry, Centre for Clinical Brain Sciences, University of Edinburgh, UK; Alzheimer Scotland Dementia Research Centre, Department of Psychology, University of Edinburgh, UK; Centre for Research on Environment, Society and Health, School of GeoSciences, University of Edinburgh, UK

## Abstract

Ambient air pollution has been associated with increased incidence of chronic disease and is estimated to contribute towards 4.2 million early deaths annually. Whilst the health impacts are well described, less is understood about the underlying biological mechanisms, particularly when considering the co-occurrence of multiple pollutants. Using an atmospheric chemistry transportation model (EMEP4UK), we generate pre-baseline sampling pollution exposure estimates for eight pollutants in Generation Scotland (N = 22,071, recruited between 2006 - 2011). Cox-proportional hazard models reveal associations between pollution exposure and all-cause dementia (PM_2.5_) and myocardial infarction (NO_3_Coarse_) over 18 years of follow-up. We perform Bayesian multivariate epigenome-wide (N = 18,512, Illumina EPIC v.1) and proteomic (N = 15,314, 133 mass-spectrometry proteins) association studies, revealing 11 pollutant-methylation associations and 140 pollutant-protein associations. We identify positive associations between exposure (PM_2.5_ and NO_3_Fine_) and epigenetic age-acceleration (PhenoAge epigenetic clock). Furthermore, we explore the development of pollutant EpiScores, assessing these in holdout and independent test sets. Our results enhance knowledge of molecular correlates of air pollution exposure, whilst providing further evidence of contributions of air pollutants to chronic disease.

## Introduction

Ambient air pollution is estimated to contribute towards 4.2 million premature deaths per year^1,2^, though estimates reach as high as 8.8 million^3^, and is associated with the incidence of several non-communicable diseases including dementia, stroke and cardiovascular disease^1,4,5^. Even low level pollution exposure has been linked to adverse health outcomes, including impaired brain health, cognitive decline and dementia risk^6–8^ and reductions in air pollution are projected to reduce mortality risk across both high and low pollution countries^9^. Air pollution is a contributor to health inequalities, as disadvantaged communities are often exposed to poorer air quality, exacerbating health disparities^10,11^. Deeper understanding of exposure-related biological processes contributing to disease will increase evidence for public health policies and reveal opportunities for intervention or treatment^12–14^.

Major systematic reviews and meta-analyses have consistently demonstrated the impact of air pollution exposure on health^15^. However, research identifying the underlying molecular pathways is more limited, if rapidly expanding in recent years^16,17^. Elucidating molecular correlates will enhance understanding of pathogenesis and has the potential to reveal exposure biomarkers for future research. However, the timescales of associated and likely dynamic, molecular changes such as DNA methylation (DNAm) or protein levels remain unclear. Epigenome-wide association studies, where the largest sample size currently reported is 8,397^18,19^, have identified associations with differential DNAm patterns in blood, implicating pathways involved in inflammation and oxidative stress^20,21^. Most of these studies focus on particulate matter (PM_10_ and PM_2.5_) and nitrogen dioxide (NO_2_), but some include other nitrogen oxides (NO_x_), ozone (O_3_), and sulphur dioxide (SO_2_)^21^, generally modelling these pollutants independently^22^. Independent modelling limits our ability to identify shared or unique contributions of individual pollutants, which may be spatially and temporally correlated^15^. Proteomic studies have implicated variation in inflammatory and coagulation proteins such as fibrinogen and CRP following both longer and shorter-term exposures^23–27^.

Here, we use an atmospheric chemistry transportation model (EMEP4UK^28^) to generate pollutant exposure estimates for eight pollutants (nitric oxide (NO), nitrogen dioxide (NO_2_), coarse nitrate (NO_3_C_), fine nitrate (NO_3_F_), ozone (O_3_), particulate matter (PM_10_ < 10µm in diameter, PM_2.5_ < 2.5 µm in diameter) and sulphur dioxide (SO_2_), see **Table 1**.), many of which have not been explored in large cohorts. This approach integrates emission data, atmospheric conditions, meteorological data and transboundary sources to estimate pollutant levels, usually for gridded surfaces^7,23,28^. After exploring nationwide trends across Scotland, we relate the pollutants to incident disease outcomes, ascertained over 18 years of follow up through data linkage to health records in Generation Scotland (GS, N = 10,739 (dementia) – 20,753 (hypertension)), and test for associations between pollution exposure and biological ageing, as measured by epigenetic clocks. Using a multivariate Bayesian regression framework, we then simultaneously study these pollution measures (averaged over 365 days prior to bio-sampling) in relation to genome-wide DNAm (N = 18,512) and proteomic patterns (N = 15,352). These analyses identify epigenetic/proteomic signals that are unique or shared across the pollutants, including two CpGs annotated to the *DIP2C* and *ATP5A1* genes and proteins, such as fibrinogen, involved in inflammation and coagulation. Together, our results provide an in-depth summary of the health and molecular correlates of pollution exposure in a Scottish cohort. Figure 1 illustrates the project overview.

**Fig. 1.**
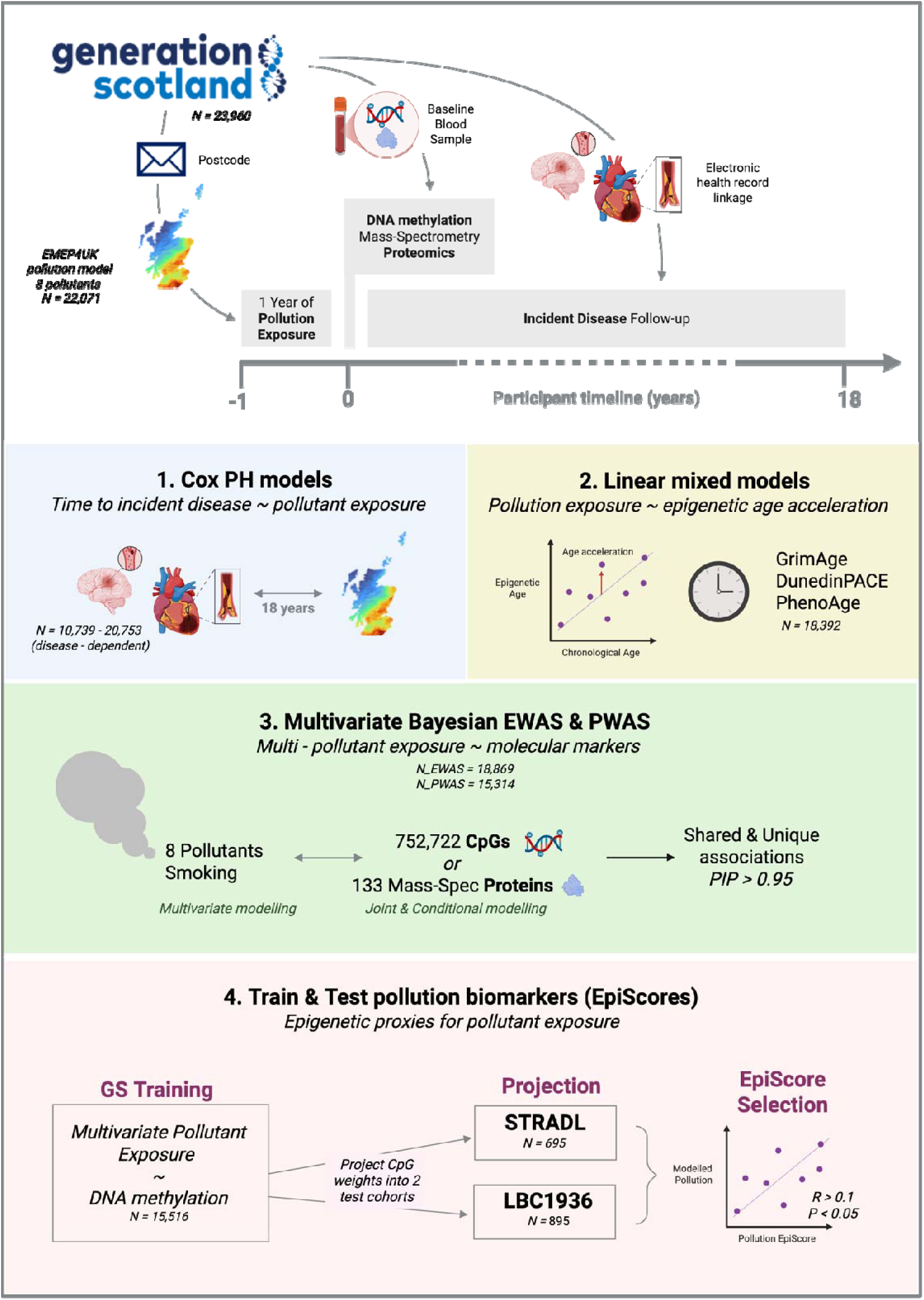
Overview of Air pollution exposure in Generation Scotland: molecular fingerprints and health outcomes. We generated pollutant exposure data for 8 pollutants in Generation Scotland using the EMEP4UK atmospheric chemistry transport model. This data was used in conjunction with blood-based DNA methylation and mass-spectrometry proteomics and incident disease outcomes over approximately 18 years of follow up for a number of analyses. 1. Cox regression was used to assess associations with incident all-cause dementia and manifestations of cardiovascular disease. 2. Linear mixed models were used to investigate associations between pollutant exposure and epigenetic age acceleration, as measured by three epigenetic clocks (GrimAgev2, DunedinPACE and PhenoAge). 3. Multivariate Bayesian modelling, where all 8 pollutants were assessed simultaneously as outcomes alongside smoking pack-years, was used to discover unique and shared associations of pollutant exposure with changes in individual molecular markers (DNA methylation – CpGs, or protein levels. 4. Using the same multivariate, Bayesian approach, we trained Pollutant EpiScores for each of the 8 pollutants in Generation Scotland. The CpG weights from the training step were projected into two test datasets: STRADL (Stratifying Resilience and Depression Longitudinally) – a sub-cohort of Generation Scotland with blood sampling approximately five years after baseline; and LBC1936 (the Lothian Birth Cohort of 1936). The pollutant EpiScores were then compared to modelled pollution in each test cohort using correlation analysis. EpiScores with a Pearson correlation of > 0.1 in both test datasets were selected as successful. Created in BioRender. Marioni, R. (2026) https://BioRender.com/s1z0zgp.

**Table 1.** Summary of pollution exposures for the 365-day time window prior to baseline appointment. See **Supplementary Table 4** and **Supplementary Figures 3** and **5** and **6** for further descriptive analysis. Mean and standard deviations were calculated for 22,071 volunteers with one-year exposure data. 24hr exceedances describes the number of individuals of the 22,071 in the GS cohort with pollution exposure exceeding the WHO 2021 AQG limit^30^ for that pollutant on more than four days based on the 365-day time window prior to their baseline appointment. This is a descriptive measure only, given these limits are defined by WHO for concentrations at fixed-site monitors. *Limits for ozone are defined differently by WHO and not included in this analysis. The remaining pollutants are not covered by the WHO guideline. Pollutants are as follows: NO = nitric oxide, NO_2_ = nitrogen dioxide, NO_3_C_ = coarse nitrate, NO_3_F_ = fine nitrate, O_3_ = ozone, PM_10_ = Particulate matter <10 µm in diameter, PM_2.5_ = particulate matter < 2.5 µm in diameter, SO_2_ = sulphur dioxide (see **Supplementary Table 27** for full definition according to the EMEP4UK model).

| Pollutant<br>[units] | Mean | Std. Dev. | WHO 2021 AQG<br>Limit [$\mu\text{g}/\text{m}^3$] | 24hr exceedances: N<br>individuals > 4 days (%) |
| --- | --- | --- | --- | --- |
| NO [$\mu\text{g}/\text{m}^3$] | 1.05 | 1.13 | - | - |
| NO<sub>2</sub> [$\mu\text{g}/\text{m}^3$] | 6.93 | 3.69 | 25 | 10,472 (47%) |
| NO<sub>3_C</sub> [$\mu\text{g}/\text{m}^3$] | 0.92 | 0.16 | - | - |
| NO<sub>3_F</sub> [$\mu\text{g}/\text{m}^3$] | 1.12 | 0.23 | - | - |
| O<sub>3</sub> [ppb]* | 31.15 | 1.97 | - | - |
| PM<sub>10</sub> [µg/m<sup>3</sup>] | 13.21 | 1.59 | 45 | 1748 (0.08%) |
| PM<sub>2.5</sub> [µg/m<sup>3</sup>] | 6.94 | 1.26 | 15 | 21,901 (99%) |
| SO<sub>2</sub> [µg/m<sup>3</sup>] | 1.69 | 0.88 | 40 | 91 (0.004%) |

## Results

### Geographical distribution of pollution

Pollution levels across Scotland during the recruitment window and one year prior (2005–2011) demonstrated marked geographical differences with higher levels concentrated around the more densely populated central belt (**Figure 2** and **Supplementary Figure 1).** There were strong spatial and temporal correlations for each pollutant, including across annual sliding time windows, with R_Pearson_ ranging from a minimum of 0.74 (NO_3_C_ for 2010 – 2011) to a maximum of 0.996 (NO_2_ for 2006-2007), **(Supplementary Figure 2** and Supplementary Tables 1 and 2).

**Fig 2.**
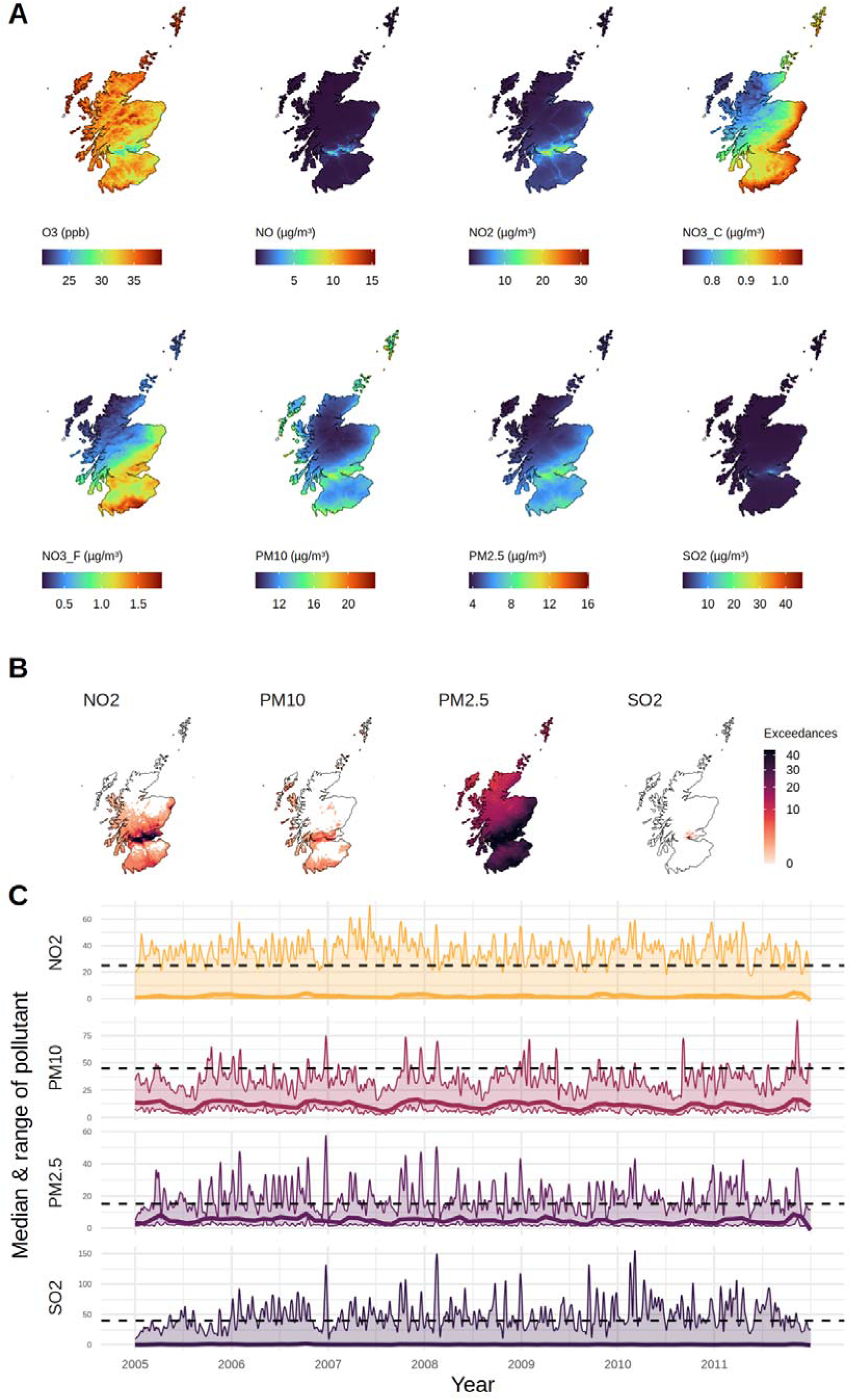
EMEP4UK pollution data across Scotland during the GS recruitment window. **A**. Average annual pollutant levels across Scotland. **B**. Number of daily exceedances of WHO limits (2021) for NO_2_, PM_10_, PM_2.5_, SO_2_ (pollutants for which 24hr limits exist) across Scotland in 2007. **C**. Daily average pollutant levels for NO_2_, PM_10_, PM_2.5_, SO_2_ showing smoothed median, maximum and minimum values and the relevant WHO 2021 limit.

Pollutant exposures for GS participants (**Table 1**, see methods), demonstrated high correlations between the 365-day average, 6-month and 7-year average (R_Pearson_>0.67) with the exception of NO_3_C_ (R_Pearson_ = 0.176, one-year: seven-year data), see **Supplementary Tables 3** and **4** and **Supplementary Figure 3**. Between-pollutant correlations demonstrate inverse correlations between O_3_ and the other pollutants, ranging from -0.75 with NO_2_, to - 0.09 with NO_3_F_,with the exception of NO_3_C_ (R _Pearson_ = 0.27; strong, positive correlations between particulate pollution (PM_10_ and PM_2.5_, R_Pearson_ = 0.92) and between particulates and NO_2_ (R_Pearson_ = 0.77 with PM_2.5_ and 0.73 with PM_10_). For full results, see **Supplementary Table 5** and **Supplementary Figure 4**. Comparing daily average individual exposures to the WHO 2021 air pollution exposure limits revealed that 47% of individuals exceeded the NO_2_ limit, and 99% of individuals exceeded the PM_2.5_ limit for more than 4 days (99^th^ percentile of daily values exceeding the limit, as defined by WHO^29^) of the 365 days assessed (**Table 1**, **Figure 1C, Supplementary Figure 5)**.

A data reduction analysis revealed that two principal components accounted for over 80% of the variance in the pollutant data (**Supplementary Table 6** and **Supplementary Figure 7**). The first principal component (PC1, contributing 58.4% of the standardised variance) demonstrated strong loadings for PM_2.5_ (0.44), NO_2_ (0.42) and PM_10_ (0.41) with positive loadings for the remaining pollutants, except for O_3_. The second principal component (PC2, contributing 24.7% of the standardised variance), demonstrated the strongest positive load for NO (0.31) with strong negative loadings for NO_3_C_ (-0.58), NO_3_F_ (-0.43) and O_3_ (-0.47).

### Pollution exposure and incident disease

Cox regression was used to explore associations between pollution exposure and health outcomes, ascertained via data linkage to health records over 18 years of follow up in Generation Scotland. We observed significant associations (P<0.004) between PM_2.5_ and incident dementia (Hazard Ratio per SD increase in exposure (HR) [95% Confidence Interval (CI)] = 1.48[1.15 – 1.9]) and between NO_3_C_ and myocardial infarction (HR[CI] = 1.79[1.25-2.56]), (**Figure 3, Supplementary Tables 7** and **8**). Our results echo findings from previous, observational studies, though to our knowledge, no study has specifically assessed long-term associations of NO_3_C_ and myocardial infarction^5,31^. Results from the analysis of associations with the first two pollutant principal components are displayed in **Supplementary Figure 8** and **Supplementary Tables 7** and **8**. These align with the individual pollutant analysis given the loadings of different pollutants onto the principal components.

**Fig 3.**
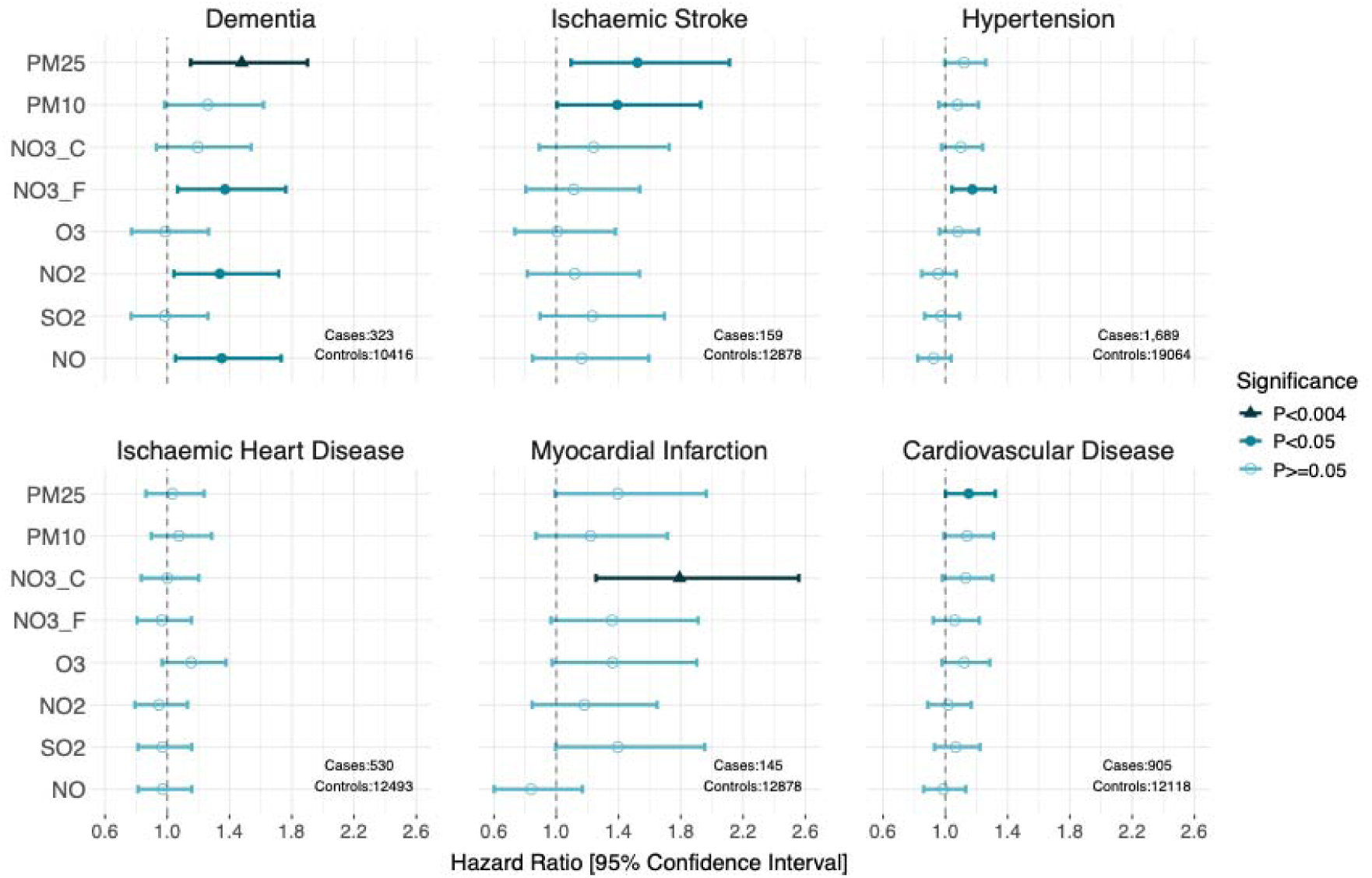
Hazard ratios from mixed-effect Cox models for models of incident disease and pollutant exposure. Results for the fully adjusted model are displayed, covariates include age, sex, BMI, smoking, alcohol units per week, SIMD rank, HDL cholesterol, Total cholesterol, a diagnosis of diabetes or hypertension (at baseline). The model for dementia additionally includes number of APOEe4 alleles and the model for hypertension does not include hypertension as a covariate. Prevalent cases for the relevant diseases are removed. Significance levels calculated as follows: P_Bonferroni_ < 0.004 (0.05/(2 x 6)) to account for the principal components of the pollutant data and number of health outcomes assessed. Pollutants are as follows: NO = nitric oxide, NO_2_ = nitrogen dioxide, NO_3_C_ = coarse nitrate, NO_3_F_ = fine nitrate, O_3_ = ozone, PM_10_ = Particulate matter < µm in diameter, PM_2.5_ = particulate matter < 2.5 µm in diameter, SO_2_ = sulphur dioxide (see **Supplementary Table 27** for full definitions according to the EMEP4UK model).

### Pollution exposure and epigenetic age

Following identification of associations with incident disease, we used linear mixed models to explore health-relevant associations at a molecular level using epigenetic clocks that track lifespan (GrimAge v.2), healthspan (PhenoAge) and pace of ageing (DunedinPACE). After adjustment for multiple testing (P_Bonferroni_ < 0.0083), PhenoAge, which captures healthspan, was associated with one-year average exposure to two pollutants NO_3_F_ (Standardised Beta = 0.02, P = 0.0014) and PM2.5 (Standardised Beta = 0.02, P = 0.0012), as shown in **Figure 4**. Full results are available in **Supplementary Table 9** and **Supplementary Figure 9**, including sensitivity analyses of different exposure-averaging windows and reduced models. This demonstrates that the addition of estimated white cell proportions has the greatest impact on the strength of associations, reducing effect sizes between model 3 (a partially adjusted model) and their addition in model 4 (fully adjusted model, see **Methods**). A 6-month averaging window demonstrated a trend towards stronger associations with PhenoAge for NO_3_F_, PM_2.5_ and NO_3_C_.

**Fig. 4.**
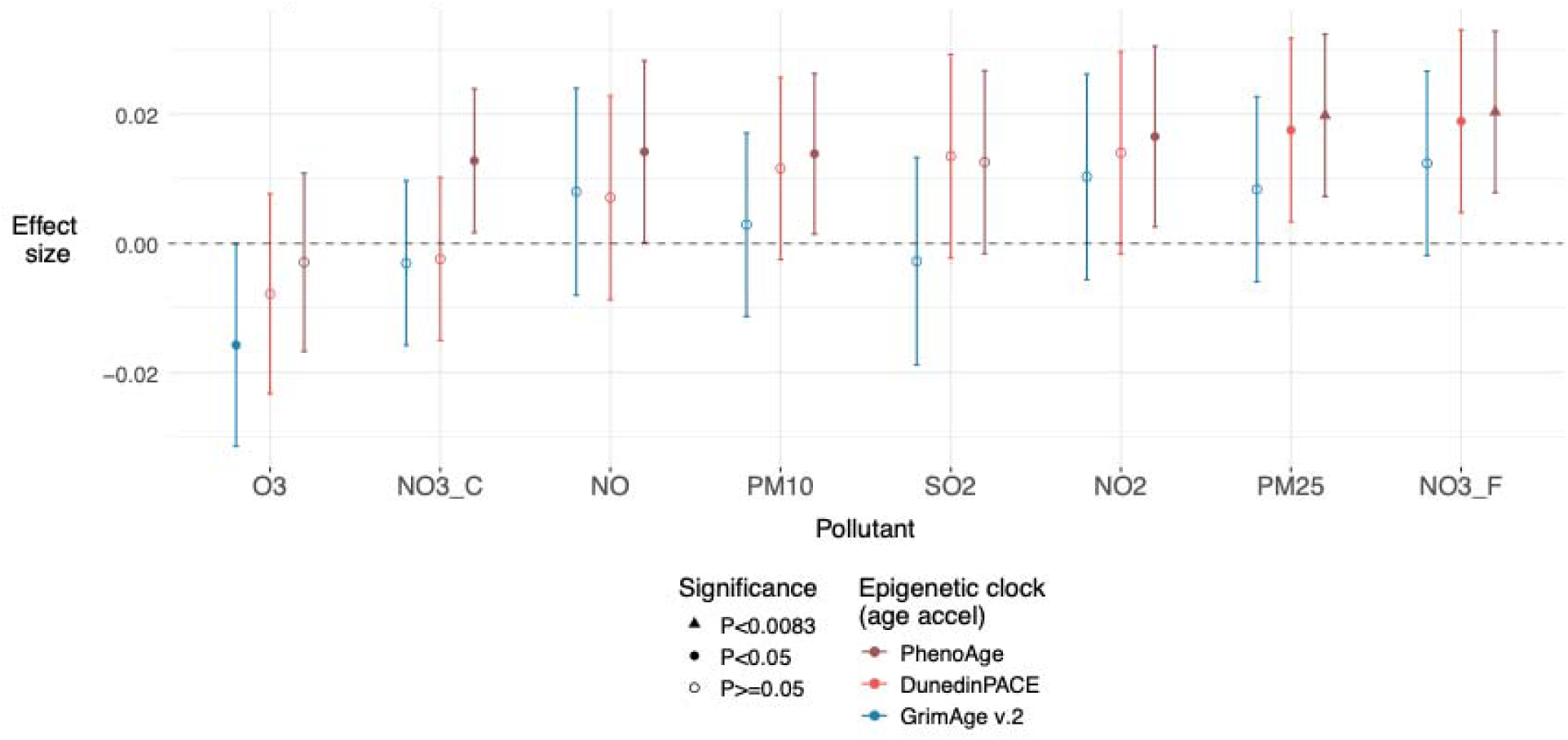
Associations between age acceleration and pollution exposure. Effect sizes (Standardised Beta) for the fully adjusted model (model 4) are displayed: Pollutant ∼ Age acceleration + age + sex + kinship + log(alcohol units) + log(BMI) + SIMD rank + log(smoking pack - years) + white cell proportions. The error bars represent 95% confidence intervals. Significance levels calculated as follows: P_Bonferroni_ < 0.0083 (0.05/(2 x 3)) to account for the principal components of the pollutant data and number of epigenetic clocks assessed. Results with nominal significance (P<0.05) are also displayed.

### Multivariate Epigenome-Wide Association Studies

To further elucidate molecular fingerprints of pollutant exposure, we ran multivariate Bayesian Epigenome-Wide Association Studies (EWAS) and Proteome-wide Association Studies (PWAS). We modelled 8 pollutants and smoking simultaneously, including smoking as an outcome given its shared route of exposure and overlap in some chemical constituents. The multivariate EWAS revealed 25 CpG-Pollutant/smoking associations, mapping to 16 unique CpGs (**Figure 5**, PIP > 0.95 and estimated effect size +/- two standard deviations, the credible interval, does not include zero). Thirteen of these 16 CpGs were annotated to 12 unique genes (Full results in **Supplementary Table 10,** String DB network (09/12/25) **Supplementary Figure 10**). Fourteen CpGs were associated solely with smoking, and the remaining two with seven and four pollutants but not smoking. The two CpGs associated with four or more pollutants were annotated to the following genes: *DIP2C* (cg26561082) and *ATP5A1* (cg16734281), with functions involved in transcription and mitochondrial ATP production respectively. If a less stringent credible interval is set (effect size +/- one standard deviation does not include zero) then some of the smoking-associated CpGs also demonstrate associations with pollutants, such as *CPAMD8* (cg15159987, NO, NO_2_, PM_10_, SO_2_) and *AHRR* (cg05575921, PM_2.5_,O_3_,NO_3_C_, NO_3_F_, PM_10_), see **Supplementary Table 11**. Variance components analyses indicated that methylation explained between 0.1% (NO) and 1% (NO_3_C_) of the variance of the pollutants and 32.0% for smoking pack years (**Supplementary Table 12, Figure 5**).

**Fig 5.**
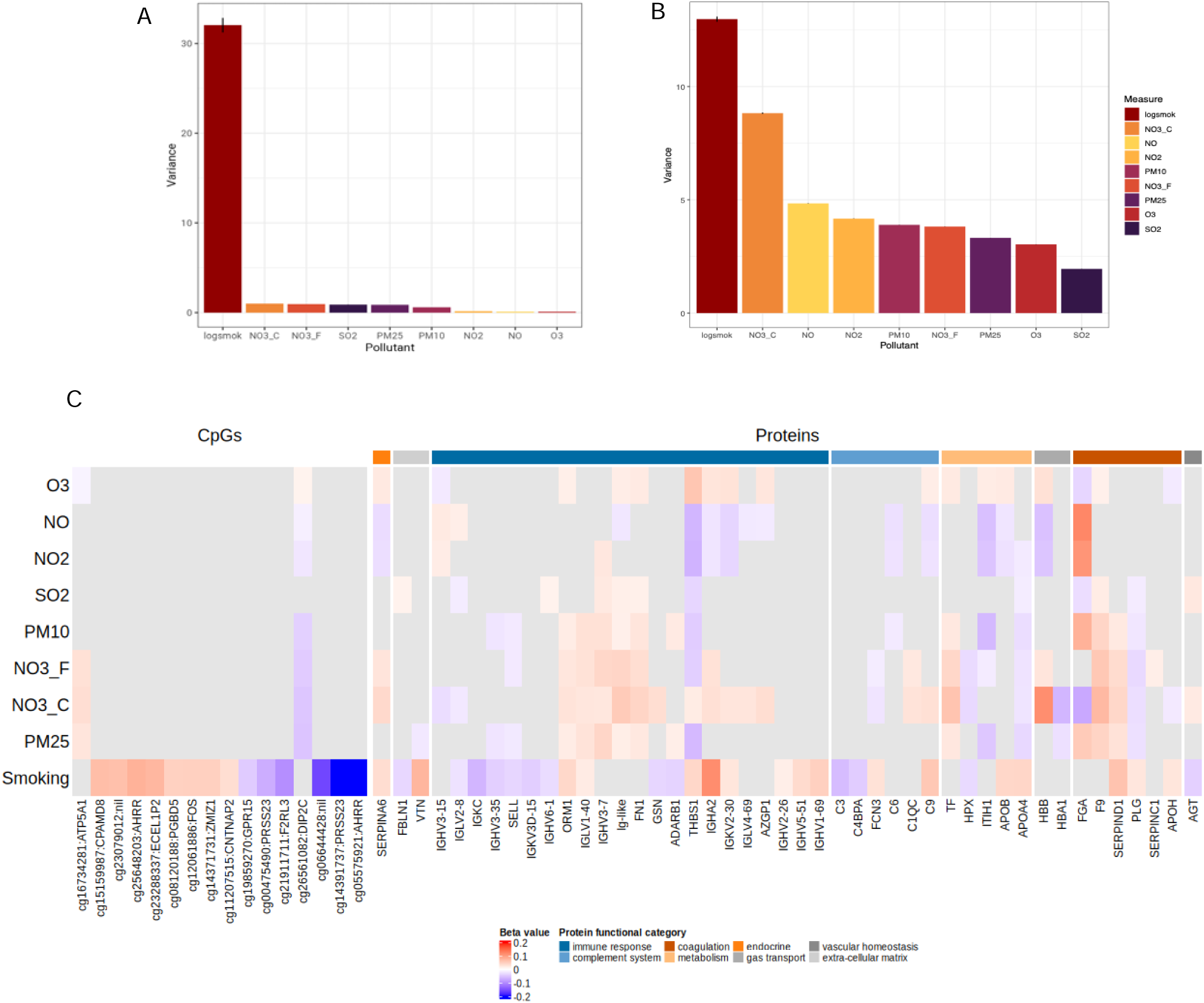
‘Omics – pollution associations identified through Multivariate Bayesian analysis. A. Variance in pollutants/smoking explained by methylation data. B. Variance in pollutants/smoking explained by proteomic data. C. Heatmap (ComplexHeatmap v. 2.24.0) displaying effect-sizes for protein-pollutant associations for proteins where at least one pollutant association was found. Where cells are grey, associations were not significant (threshold for significance: PIP > 0.95 and 95% credible interval which does not cross zero) for that protein-pollutant pair or CpG-pollutant. Protein functional category was determined using Gene Ontology via UniProt^33^. MultivAriate Joint bAyesian model (MAJA) was used to perform the analysis (see **Methods**).

### GMRM EWAS

Sensitivity analyses, using univariate Bayesian EWASs of each of the eight pollutants identified the same *DIP2C* CpG as highlighted by the multivariate model. However, the *ATP5A1* association was unique to the multivariate approach (**Supplementary Table 14**). Replicated associations (N = 15) between univariate and multivariate approaches demonstrated highly correlated effect sizes (R_Pearson_ = 0.998), see **Supplementary Figure 11**. Univariate analyses using 6-month and 7-year exposure data identified seven and one CpG-pollutant associations, respectively. The association between cg26561082 (*DIP2C*), and PM_2.5_ was also identified in the univariate EWASs with 6-month average exposure (**Supplementary Figure 11)**.

### EWAS Catalog replication

We next characterised our EWAS findings through look-up of the EWAS Catalog. Of the 16 CpGs we identified, 15 have entries in the fully-filtered EWAS Catalog (N > 1000, P < 3.6 x 10^-8^, sample type = whole blood, see methods). Eleven of these have previously been reported in association with smoking (**Supplementary Table 11)**. We additionally identified two previously un-reported CpG – smoking associations: cg08120188 (*PGBD5*) and cg12061886 (*FOS*). Further, cg23288337 (*ECEL1P2*) has only recently been reported in a Bayesian EWAS of smoking and in a meta-analysis of smoking^32^. Other traits associated with the 15 CpGs in the EWAS catalogue included body mass index, alcohol consumption, a variety of lung function tests including (FEV1, FVC, FEV1/FVC, DLCO/VA) and incident health conditions including lung cancer, ischaemic heart disease, type 2 diabetes, COPD and CKD (for full list see **Supplementary 11)**. None of the CpGs have previously been associated with air pollutant exposure. Of the two pollutant - associated CpGs, cg26561082 (*DIP2C*) has previously been associated with age. No previous associations were identified for cg16734281 (*ATP5A*1). Enrichment analysis did not identify pathway-enrichment in probe-associated genes from our results (**Supplementary Table 15**).

### Multivariate PWAS

Multivariate Bayesian regression modelling revealed 170 protein-pollution associations (PIP > 0.95) between 45 proteins and all outcome traits, with some proteins exhibiting associations across multiple traits (**Figure 5**). One protein (Complement C3) associated with smoking and no air pollutants. Several immunoglobulin components (e.g. heavy variable, lambda variable) demonstrated associations with multiple pollutants in both positive and negative directions. We additionally identified several proteins involved in coagulation which demonstrate associations with multiple pollutants. Full results are available in **Supplementary Tables 16and 17**. From the variance components analyses, the proteome-wide data explained between 1.95% (SO_2_) and 8.82% (NO_3_C_) of the variance in pollution exposure and 12.97% in the variance in smoking pack-years (**Figure 5**, **Supplementary Table 18**).

### GMRM PWAS

Univariate Bayesian PWASs in GS, using the 1-year exposure data identified 189 protein – pollutant or smoking associations (PIP > 0.95); 41 with smoking and 148 with the pollutants. Replicated associations between univariate and multivariate approaches (N = 127 including smoking) demonstrated highly correlated effect sizes (R_Pearson_ = 0.99). Univariate analyses using 6-month and 7-year exposure data identified 105 protein-pollutant for each time duration. (**Supplementary Tables 19 - 21** and **Supplementary Figure 12)**.

We compared our results with those from a recent UK Biobank analysis which investigated proteins associated with exposure to PM_2.5_ and NO_2_ in 51,357 participants^23^. This study used multiple linear regression models to investigate associations between 2,265 proteins (Olink Explore platform) and PM_2.5_ and NO_2_, discovering 138 associations with PM_2.5_ and 143 with NO_2_ (FDR < 0.05). There is an overlap of 57 proteins between the two datasets. However, of the 26 proteins found to be associated with PM_2.5_ or NO_2_ in our results, none had significant results in the comparator study, and, correspondingly, none of the proteins identified in the comparator study had significant associations with any pollutant in our results. This likely reflects substantial methodological differences between the studies including in the approach to pollutant exposure modelling and association testing (see **Discussion**). Enrichment analysis via PANTHER did not identify any enriched pathways (**Supplementary Table 22)**.

### Pollution EpiScore

Broadly analogous to polygenic scores in GWAS, we generated DNAm EpiScores for each pollutant using multivariate EWAS in GS. In a holdout test set of GS and the external LBC1936 cohort, there was weak evidence for associations between the EpiScore and its corresponding measured pollutant – only the PM_10_ EpiScore showed a significant association in the GS holdout test set (R_pearson_=0.12, P=0.002). The full results are presented in **Supplementary Tables 23and 24**.

## Discussion

We performed a large-scale multi-omic analysis of eight measures of pollution exposure. We demonstrated associations with common incident disease outcomes in addition to accelerated biological ageing. We identified epigenetic and proteomic signals that were both shared and unique to pollutants and smoking. Specifically, we highlighted a single CpG locus, mapping to *DIP2C* that associated with seven of eight pollutants, and proteins linked to inflammation and coagulation that associated differentially with multiple pollutants.

The majority of studies assessing air pollution biomarkers thus far have focussed on two to three pollutants; most commonly PM_2.5_, NO_x_, NO_2_ and PM_10_^15^. We expand this to include additional pollutants which may have differing and/or synergistic biological effects and are often spatially and temporally correlated with more frequently studied pollutants^34^.

Air pollution was assessed from 2005-2011 in Scotland, demonstrating comparatively low air pollution^35^. However, for PM_2.5_ and NO_2_, these exposures exceed WHO air quality guidelines (2021) in 99% and 47% of individuals assessed, respectively, albeit with marked geographical variation^30^. Many of the pollutants are highly correlated (**Supplementary Table 5., Supplementary Figure. 4**). This is supported by principal component analysis where the first two components contribute to 83.2% of the overall standardised variance. Within these two principal components, several pollutants demonstrate similar loadings indicating high co-variation amongst the pollutants modelled. For example, the similar contributions of PM_2.5_, NO_2_ and PM_10_ to PC1 (with loadings of 0.44, 0.42 and 0.41, respectively), suggests comparable contributions to the dominant pollutant pattern. This could be consistent with a shared source such as transport or industrial-related combustion. PC2, with strong loadings for NO_3_C_, NO_3_F_ and O_3_, could reflect secondary atmospheric chemistry processes related to photochemical oxidation. Therefore, independent modelling of pollutants in the context of EWAS, for example, may miss important contributions of co-varying pollutants. Our multivariate approach helps leverage the co-variation of pollutants to identify associations which are common or unique to pollutants which may have differing or synergistic effects.

We used a widely recognised and applied atmospheric chemistry model to estimate pollution exposure for eight pollutants based on home address. This model supplies estimated daily averages at a spatial resolution of 3km by 3km and is robustly benchmarked against monitored pollutant levels where available^36^. It provides an estimate which considers a wider-range of pollutant sources than traditional land-use regression models and is more robust to errors that can be introduced by higher-spatial resolution land-use regression modelling^37^. Individual exposure estimates can then be calculated based on residential postcode, enabling exposures to be estimated at scale, allowing for the limitation that this approach does not capture indoor, travel or occupation – related exposures^38^. For modelling purposes, we have used averaged estimated exposure for the 365 days prior to blood sampling, with 6-month averaged exposure and a 7-year averaged exposure as comparators. Previous work has suggested that the methylation signature of smoking is time and dose dependent^39^ and therefore we anticipated that short and longer-term methylation signatures may differ depending on changing exposures. The 365-day time frame accounts for shorter-term fluctuations in pollutant levels due to seasonal and meteorological affects which may be less relevant to longer term health outcomes. The use of comparator time frames allows the exploration of whether molecular changes more closely reflect nearer term or longer-term exposures.

Numerous, large studies have identified associations of ambient air pollution exposure and negative health outcomes, including cardiovascular disease, dementia, asthma and lung conditions^12,40,41^. Though, smaller in size than many of these larger studies, we identified associations with dementia and cardiovascular-related diseases in the Generation Scotland cohort, reinforcing the growing evidence that even low-levels of pollution exposure can have negative health impacts.

We also explored associations of pollutant exposure with epigenetic age-acceleration calculated by epigenetic clocks modelled on healthspan, lifespan and pace of ageing. Mixed-effect models identified positive associations between one-year pre-sample pollution exposure and epigenetic age-acceleration according to the PhenoAge epigenetic clock. Though effect sizes are small per standard deviation, if we considered PM_2.5_ concentrations in Lahore, Pakistan, currently rated the most polluted city in the world (Air quality index AQI^+^, 28/10/2025), with an average annual concentration of 102.1 μg/m^3^ in 2024^42^ compared to 6.9 μg/m^3^ (S.D. 1.26 μg/m^3^) for GS participants, the associated deviation in biological age could be expected to be proportionally larger, though this relationship may not be linear. That the inclusion of white cell proportions in model 4 (see **Methods**) demonstrates the strongest attenuation of associations between pollutants and DunedinPace_EAA_ or GrimAge2_EAA_ may reflect that neither include white cells in their algorithm, in comparison to PhenoAge which does (**Supplementary Figure 9B**). It is possible that the trend towards stronger associations for 6-month average pollution exposure and PhenoAge_EAA_ but a 7-year averaging period for GrimAgev.2_EAA_ (**Supplementary Figure 9A**) may reflect the heterogeneity in longitudinal variation of methylation at different sites in response to stimuli and the differential inclusion of methylation sites for each clock. However, this would need further experimental evidence to be fully explored.

Our multivariate analysis of 1-year pre-sample average exposure identified 11 CpG – pollutant associations between two CpGs and seven of eight pollutants. One of these CpGs (cg26561082) was negatively associated with six pollutants and demonstrated a positive association with O_3_. It demonstrated no association with smoking. *DIP2C* is a relatively under-characterised gene, however, it is highly conserved across invertebrates and vertebrates, and widely expressed across tissues^43,44^. It has proposed roles in neuro-developmental disorders, metabolism and tumour suppression^45^. Further, as the gene contains a *DMAP1* binding domain, it has been suggested to play a role in *DNMT1*-dependent methylation^46^. In EWAS studies, differential methylation at this site (in studies with N > 1000) has previously been associated with age and Idiopathic/heritable pulmonary arterial hypertension^47,48^. Methylation at cg16734281 (*ATP5A1*) was positively associated with PM_2.5_, NO_3_C_, NO_3_F_ and negatively associated with O3 exposure. *ATP5A1* encodes a subunit of ATP synthase, a crucial enzyme in mitochondrial ATP production^49^. Aberrant expression of *ATP5A1* has been associated with cancer including renal cell carcinoma and glioblastoma^50,51^. Additionally, we identified two, to our knowledge, previously unidentified CpG sites associated with smoking. These were cg08120188 (*PGBD5*) and cg12061886 (*FOS*). Overall, the number of EWAS associations for smoking (N_CpGs_ = 14) was greater than those for the pollutants with the most associations (N_CpGs_ = 2). This is unsurprising, given the direct route of exposure and higher concentration of particulates and chemicals in smoking inhalation compared to that of ambient air pollution.

The univariate analyses demonstrate the added benefit of multivariate analyses of low-variance correlated traits such as pollution exposure. For the directly comparable analysis of the 1-year data, many fewer associations were identified for the low-variance pollutants in the univariate analysis (6 versus 11). Despite the increased power to detect associations in the multivariate regression analysis, there was minimal evidence to suggest that DNAm EpiScores can act as surrogates for pollution exposure using a training sample of this size in a relatively low exposure setting.

Analysis of the high-abundance proteome as measured by mass-spectrometry identified 170 protein – pollutant/smoking associations for 45 unique proteins (**Figure 4**). When broadly categorised according to gene ontology information from UniProt^33^, 28 of the associated proteins were immune-related, with 6 of these functioning in the complement system and a further 6 were involved in coagulation pathways. The remaining 11 are involved in metabolic and endocrine pathways, gas transport, vascular homeostasis or play a role in the extra-cellular matrix (**Figure 4**). Assessment for enrichment via PANTHER^52^, did not identify any enriched pathways (**Supplementary Table 22**). Notable examples include Thrombospondin-1 (TSP-1) which demonstrated negative associations with seven of eight pollutants, and positive associations with NO_3_C_ and smoking. TSP-1 is a matricellular protein and has multiple roles related to cell proliferation and migration in the context of inflammation and angiogenesis. Decreased TSP-1 protein expression is associated with progression in several cancers, whilst circulating levels can be increased in people with cancer^53^. The fibrinogen alpha-chain demonstrated associations with seven pollutants (all except NO_3_F_) and smoking. It is one of three subunits of fibrinogen, encoded by three genes (*FGA, FBG, FGC).* Fibrinogen is a precursor to insoluble fibrin – a critical component of blood clots, is increased in inflammatory states and also has roles in antimicrobial response^54^. Further associations between multiple pollutants and coagulative proteins include Factor IX (F9), Heparin Co-Factor 2 (SERPIND1) and Plasminogen (PLG); and between multiple pollutants and inflammatory/immune proteins include multiple immunoglobulin components, L-Selectin (SELL) and Gelsolin (GSN). These associations, though reflective of the high-abundance proteome as measured via mass-spectrometry, are supported by previous work identifying changes in inflammatory proteins^55^.

The lack of overlap between our analysis and the UK Biobank proteomics analysis of PM_2.5_ and NO_2_ may be due to several factors: different methods of protein measurement - mass spectrometry versus Olink; different approaches to pollution modelling - atmospheric chemistry transport model versus land use regression and 365 day average prior to blood sample versus yearly averages based on available data; and finally different modelling approaches – a multivariate Bayesian approach with joint and conditional modelling of molecular markers versus multiple linear regression modelling.

Our study has some limitations. Firstly, we relied on home postcode to generate exposures which may not fully reflect occupational, travel-associated or indoor exposures and assumes residential stability in the preceding year. Previous studies have identified that this may lead to an underestimation of exposure estimates and therefore downstream health effects by 12-16% ^56,57^. The use of a modelling approach allows retrospective, high coverage and high-resolution data which can be practically applied to such a large cohort, allowing us to look at outcomes now for exposures in the past. However, individualised, monitored exposure would generate the most accurate data and, to assess similar health outcomes, would require prospective follow-up over a similar timeframe. Secondly, we currently do not have sufficient evidence to know what time frames are most pertinent to health-relevant molecular changes and thus we have chosen these time frames as representative exposures – similar to approaches used in previous studies^23,58,59^. It is possible that exposures at particular ‘vulnerable’ periods of life, such as during the neonatal period or middle adulthood may have more profound effects^58^, however, we have not addressed this question in our study. Longitudinal studies with repeated measurements and personalised pollution exposure monitoring would be a preferred, if logistically and technically challenging, approach. As wearable device technologies advance, it is possible that such an approach will become more feasible in the future.

Finally, Generation Scotland is currently one of the largest cohort studies of its kind with both DNAm, proteomic data and health-record linkage which provides a unique opportunity to explore the molecular impacts of such a challenging phenotype. However, the cohort is of predominantly white European ancestry and Scotland itself is a country with relatively low ambient air pollution compared to others^12^. Therefore, these results may not be generalisable to other populations, and similar studies should explore more diverse ancestries and geographical areas where pollution exposure may be higher.

The disease analyses were limited by the use of secondary care data, with the exception of dementia diagnosis, as the basis for prevalent and incident diagnoses which may underestimate the number of cases. Further, we use broad definitions of disease when considering dementia (all-cause) which may obscure any subtype specific effects. This is evident when we consider diagnoses contributing towards our broader cardiovascular disease definition, revealing significant associations with myocardial infarction.

## Conclusions

We have generated pollution exposure data for Generation Scotland, one of the largest cohorts currently available with health record-linkage and multi-omic measurements including methylation and proteomics. Analysis of this data has revealed expected associations between increased pollutant exposure and negative health outcomes, increased biological age acceleration as measured by the PhenoAge epigenetic clock capturing healthspan and several epigenetic and proteomic markers associated with exposure. We were able to generate a pollution EpiScore for PM_10_ which met our performance threshold in one of our two test-sets. That the majority of the EpiScores did not meet our threshold for further investigation is unsurprising, given the low variance explained by methylation data.

Our EWAS results demonstrate a greater number and strength of associations with smoking compared to air pollution exposure. This reinforces our current understanding of the severe and wide-ranging impacts of smoking on health. However, the ubiquitous and near-constant exposure to air pollution is challenging to modify at an individual level, and our results including associations with health outcomes despite comparatively low exposure, demonstrate that it remains a threat to public health. Emerging sources impacting air quality in both Scotland and other countries globally include the rise in domestic use of wood-burning stoves and wildfires^60,61^. There are many countries and cities where air pollution remains at very high levels with evident immediate health impacts^12,62,63^. Our study highlights the danger of longer-term exposures, even at what may be considered relatively low levels of exposure, whilst also revealing previously unidentified molecular markers of both pollutant and smoking exposure using a Bayesian multivariate method.

## Methods

### Generation Scotland

Generation Scotland is an epidemiological study with genetic, DNA methylation, clinical including health-care record linkage, and socio-demographic data for approximately 24,000 volunteers, in approximately 5,500 family groups^64^, from across Scotland, aged 17-99^65^. Recruitment occurred between 2006 and 2011, and blood samples used for omics profiling were taken during the clinic visit, alongside health, cognitive and lifestyle questionnaires. Electronic-health records linkage to secondary and primary care data allows analysis of prevalent and incident disease^66^. Demographics are available in **Supplementary Table 25**. STRADL is a subset of 1188 individuals from the original GS cohort with additional assessments and blood samples approximately 5 years after the study baseline^67^.

Participants provided written informed consent. Ethical approval was received from the NHS Tayside Committee on Medical Research Ethics (REC Reference Number: 05/S1401/89) and Research Tissue Bank status was granted by the East of Scotland Research Ethics Service (REC Reference Number: 20/ES/0021).

### DNA Methylation in Generation Scotland

DNA methylation (DNAm) was profiled from sodium bisulphite treated DNA from whole-blood baseline samples on the Illumina Infinium HumanMethylationEPIC BeadChip array v1.0^68^. Sample processing and quality control (QC) of methylation data has previously been described in full^69^. A total of 752,722 CpG sites were included following quality control (see **Supplementary Methods 1)**. For downstream analysis, methylation data were normalised using the dasen method (R package wateRmelon v.2.2.0^70^). Methylation M-values were used throughout and were calculated using the beta2M function in the R package lumi v.2.30.0^71^. M-values are the log2 ratio of intensities of methylated to unmethylated probe and are recommended for differential methylation analysis^72^. DNAm in STRADL was processed in the same way.

### Mass spectrometry proteomics in Generation Scotland

Full details of the protocol have been described previously^73^. Briefly, serum samples were pre-processed for protein denaturation and trypsinisation, prior to liquid chromatography - mass spectrometry (LC)-MS, using the Agilent 1290 Infinity II system and TripleTOF 6600 mass spectrometer (SCIEX) and a scanning SWATH method^74^. Output data were processed by DIA-NN^75^, identified using a spectral library^76^ with precursor false discovery rate (FDR) set to 1%. R was used for further post-processing including within-batch drift and between-batch correction^77,78^. Identified proteins and peptides were mapped to Universal Protein Resource (UniProt) IDs^33^. 133 of these were uniquely mapped to one protein and these proteins were used for downstream analysis (full annotations in **Supplementary Table 26**). At the time of analysis, data for 15,818 GS participants had been generated.

### Lothian Birth Cohort 1936

The Lothian Birth Cohort 1936 (LBC1936) is a longitudinal volunteer study. Individuals who were born in 1936 and undertook the Moray House Test No.12 aged ∼11 years whilst at school in Scotland were invited to join the LBC1936 aged ∼70 years^79^. Recruitment occurred between 2004-2007, with participants predominantly residing in the Edinburgh and Lothians region of Scotland. Participants undergo follow-up triennially, including physical, cognitive and medical assessments^80^. Data from wave 1 (mean age 69.5 years, 50% female) were used for this analysis (N = 1091).

All participants gave written informed consent. Ethical approval for LBC1936 study was obtained from the Lothian Research Ethics committee (LREC/2003/2/29) and Multi-Centre Research Ethics Committee for Scotland (MREC/01/0/56).

### DNA methylation in LBC1936

DNA was extracted from whole blood samples and DNA methylation measured via the Illumina 450K methylation array at the Edinburgh Clinical Research Facility. Quality control steps have been described previously^81^. Briefly, internal controls were used for background correction and raw-intensity normalisation. Manual inspection was carried out to remove low quality samples secondary to inadequate hybridisation, nucleotide extension, issues related to bisulphite conversion or staining signal. Samples with a mismatch in genotype and SNP control probes or in reported and DNA methylation-predicted sex were removed. Finally, samples with a low call rate (< 450,000 probes detected at P < 0.01) and probes with a low detection rate (< 95% at P < 0.01) were removed.

### Pollution exposure (EMEP4UK)

We used daily average low-altitude ambient air pollution data from the EMEP4UK (rv4.36)^28^, an atmospheric chemistry transport model, based on the EMEP MSC-W model^82^ in tandem with meterological input data from the Weather Research Forecast model (v4.2.2)^83^. This model has been described in full previously^84–86^. Briefly, it uses local and transboundary emissions data, with meteorological input to generate pollutant concentrations at a spatial resolution of 3km^2^ for the UK. The data are evaluated using the UK Automatic Urban and Rural Network (AURN) monitoring stations.

Scotland-wide data were interrogated to explore spatial and numerical distributions of means and ranges and year to year correlations and aggregate year correlations for the GS recruitment window and one year prior, 2005 – 2011 (Terra package v 1.8.29^87^)). The following pollutants were considered: particulate matter with diameters < 10 and 2.5 µm (PM_10_, PM_2.5_), coarse and fine particulate nitrates (NO_3_F_, NO_3_C_), sulphur dioxide (SO_2_), ozone (O_3_), nitrogen dioxide (NO_2_) and nitrogen oxide (NO). The full definition of each pollutant as per the EMEP4UK model can be found in **Supplementary Table 27.**

Pollution exposure was assigned to Generation Scotland participants (N = 22,071) using home postcode at the time of baseline appointment (occurring across 2006 - 2011), using the PostcodesioR^88^ (v. 0.3.1) and Terra packages^87^ (v1.8.29) in R, version 4.1.3. A one-year (365-day) time window preceding the baseline appointment was used to extract daily mean exposures for each participant and calculate a mean 365-day exposure. As comparators, mean 6-month (183-day) exposure and 7-year exposure (the recruitment window and one-year prior 2005 – 2011 inclusive), were also calculated. Annual and 24-hour exceedances of the WHO 2021 air quality guideline limits^30^ were calculated for PM_2.5_, PM_10_, NO_2_ and SO_2_, by individual. Averaged pollutant data were scaled to a mean of 0 and unit variance for analysis. Principal components analysis was run on the 365-day average data in R using PCAtools^89^ (v. 2.12.0). Using the same methodology as for Generation Scotland, pollution exposure was assigned to LBC1936 participants in wave 1 (N = 1,088) using home postcode at the time of baseline appointment (occurring across 2004 – 2007) for a 365-day time window preceding blood-draw appointment. Similarly, using the same methods, exposure was identified for STRADL participants in the 365 days prior to blood-sampling (occurring approximately 5 years after baseline GS sampling).

### Pollution exposure and Health outcomes: Cox Proportional Hazard models

To explore whether our data supports previously identified associations between pollution exposure and health outcomes we undertook Cox proportional hazard modelling^90^ for pollution exposure and incident disease with a maximum follow up of 18.6 years (dementia models) or 17.7 years (remaining disease models). Diseases considered included: all-cause dementia, hypertension, and a broad, inclusive definition of cardiovascular disease which incorporates diagnoses including myocardial infarction, ischaemic stroke, ischaemic heart disease and any death relating to cardiovascular disease. We additionally explored individual outcomes contributing to the broad cardiovascular disease definition: myocardial infarction, ischaemic stroke, and ischaemic heart disease to explore any subtype-specific effects. Diagnoses were established from secondary care records, using CALIBER/HDRUK consensus definitions^91^ and cardiovascular disease-related death was determined using ICD codes I00-99, in line with previous work on cardiovascular disease carried out in Generation Scotland^92^. All-cause dementia cases were determined from both primary and secondary care records. For full details of disease definitions, see **Supplementary Methods 2** and **Supplementary Table 28**.

Three, iterative mixed-effects models were run for each disease – pollutant pair. The basic model adjusts for age, sex and kinship. Kinship was adjusted for using a kinship matrix constructed using the R package kinship2 (v.1.9.6) as specified above. The second additionally adjusting for smoking pack-years (one smoking pack year equates to twenty cigarettes smoked daily over one year), body mass index (BMI = weight in kg/height in m^2^), alcohol consumption (self-reported units/week), Scottish Index of Multiple Deprivation rank (SIMD), 2009^93,94^ and, in the case of the dementia model, APOEe4 allele count. The final model adds adjustment for high density lipoprotein cholesterol (HDL) and total cholesterol and a diagnosis of diabetes or hypertension (excluding the hypertension model) at baseline. Missing covariate data were imputed using kNN (k=5, VIM v. 6.6.2) and prevalent cases for the relevant disease were removed from the dataset (see **Supplementary Methods 2**). Models were run using the survival^95^ and coxme packages^96^. We also explored associations using the first two principal components of the pollution data, in place of the individual pollutants for one-year pollution exposure, using the fully adjusted model. Full details of data-preparation for each disease, and model checks, including proportional hazards, are included in **Supplementary Methods 2** and **Supplementary Figures 13** and **14**. P_Bonferroni_ was set at 0.004 (0.05/(2 x 6) using the number of principal components which explain >80% of the variance in the pollutant data (two) and the number of diseases compared (**Supplementary Table 6).**

### Epigenetic Age acceleration and Pollution exposure

We explored whether pollution exposure was associated with epigenetic age-acceleration according to three clocks that track lifespan, healthspan and pace of ageing: PhenoAge^97^, GrimAge version 2^98^ and DunedinPACE^99^. Epigenetic age for Generation Scotland participants with methylation data (N = 18,869) was derived using the biolearn platform^100^. Epigenetic age acceleration was calculated by taking the residuals of the model Clock Age ∼ chronological age. Residuals were scaled to a mean of 0 and unit variance for downstream analysis. We ran a series of iterative mixed-effects models for each of the 8 pollutant-exposures from the EMEP4UK model^28^. Kinship was included as a kinship matrix constructed using the R package kinship2 (v.1.9.6). This incorporates maternal and paternal identifiers for each participant in the cohort as a matrix such that values of 0.5 are specified for parent-offspring or sibling pairs. Additional covariates included alcohol consumption (units/week), smoking pack-years, BMI, SIMD rank and six Houseman-estimated^101^ white cell proportions (CD4T, CD8T, Bcell, Monocytes, Natural Killer cells and Eosinophils, neutrophils were dropped due to multi-collinearity). Covariates were log-transformed where necessary to account for skew, with a constant of 1 added to units of alcohol per week and smoking pack-years to account for those with no alcohol or smoking intake. Complete pollutant and age-acceleration data was available for 18,556 participants. After filtering to BMI >=16 and <=50, to remove extreme outliers, 18,392 samples remained for analysis. Demographic details are available in **Supplementary Table 28** and correlation analysis between the pollutants and included covariates in **Supplementary Figure 15**. Missing data for the covariates (alcohol units per week = 1663, smoking pack-years = 384, SIMD rank = 560) were imputed using kNN (k = 5) from the R package VIM (v. 6.6.2)^102^. Associations are detailed for p-value cut-offs of < 0.05 (nominal), and < 0.0083 (0.05/(2×3)) to account for the pollutants (first two principal components) and number of Epigenetic clock comparisons. Iterative models were run as outlined below:

**Model 1:** pollutant ∼ age acceleration + age + sex + kinship
**Model 2**: pollutant ∼ age acceleration + age + sex + kinship + alcohol units + BMI + SIMD rank
**Model 3**: pollutant ∼ age acceleration + age + sex + kinship + alcohol units + BMI + SIMD rank + smoking
**Model 4**: pollutant ∼ age acceleration + age + sex + kinship + alcohol units + BMI + SIMD rank + smoking + white cell proportions

### EWAS of air pollution exposure

We undertook multivariate (MAJA^103^) and univariate (GMRM^104^) Bayesian epigenome-wide association analyses of air pollution and smoking behaviour. 18,869 individuals from Generation Scotland had complete pollutant and methylation data. These were filtered to individuals with BMI >=16 and <=50 (N_remaining_ = 18,512). In the multivariate analyses, smoking pack-years was included as an outcome variable due to shared exposure route and constituents with pollutants^105,106^. This allows identification of pollutant-marker associations shared with smoking. The smoking variable was log-transformed (+1 to account for those with 0 pack-years) due to skew, scaled to a mean of zero and unit variance prior to the EWASs and missing data (N= 391) imputed via kNN from VIM (v. 6.6.2)^102^. Methylation M-values were pre-regressed for covariates using the limma package (version 3.60.4) in R^78^. Covariates with known impacts on DNAm or potential associations with pollution exposure were included and log-transformations were applied where necessary due to skewed distributions. Included covariates were age, sex, measurement batch, log transformation of BMI, log transformation of alcohol units consumed per week, and SIMD rank 2009. Prior to regression, covariates were scaled (mean of 0, S.D. of 1) and any missing data were imputed using kNN function from VIM (v. 6.6.2)^102^, (N_missing_ BMI = 120, SIMD rank = 564, Alcohol units = 1701). Residuals from the output of a linear model for each CpG were scaled to have a mean of zero and unit variance prior to the EWASs. All analyses outside of MAJA and GMRMomi were conducted in R version R 4.4.2. CpG sites were annotated using the minfi package in R (IlluminaHumanMethylationEPICanno.ilm10b4.hg19), version 1.50.0^107^, to establish chromosome, probe position, relation to CpG-island and any nearby genes.

### Multivariate approach: MAJA

A MultivAriate Joint bAyesian model (MAJA)^103^ EWAS was performed to identify CpG – pollution associations unique and shared for each pollutant. This approach simultaneously models all eight pollutant exposures from the EMEP4UK model and smoking pack-years as outcomes. Prior distributions of effect sizes are assumed to have a multivariate spike and slab distribution to allow for markers with zero effect. Input markers are modelled jointly and conditionally, implicitly controlling for unknown variables such as white-cell proportions and genetic architecture. 1500 iterations were run, with 500 ‘burn-in’ iterations discarded prior to averaging effect sizes over the last 1000 posterior samples. Significant associations were taken to be those with a posterior inclusion probability (PIP) of greater than 0.95. Due to the multivariate normal prior, CpG probes are included in the model if at least one of the traits has a non-zero effect size. To check if a probe is associated with a certain trait, we filtered to a posterior inclusion probability (PIP) if greater than 0.95 and to probes with an effect size where the credible interval (+/- two standard deviations) does not cross 0.

### Univariate approach: GMRM EWAS

For comparison, univariate Bayesian EWASs of pollution exposure were performed for each pollutant and smoking in turn using GMRM-omi^104^. Methylation and pollutant data were prepared as above. GMRMomi is an implementation of Bayesian penalised regression^104^ based on a genomics data framework and adapted for large-scale multi-omics data. Similarly to MAJA, it models all CpGs jointly and conditionally, implicitly controlling for unknown variables as above. GMRM uses Gibbs sampling to draw from posterior distributions. Prior mixture variance proportions were set to 0.0, 0.001, 0.01, and 0.1, equivalent to negligible, small, medium and large CpG effect sizes. 2000 model iterations were run with 750 ‘burn-in’ iterations discarded prior to averaging effect sizes over the last 1250 posterior samples. As a sensitivity analysis, univariate Bayesian EWASs were also run for the eight pollutants for average exposure over six months and seven years. Significant associations were taken to be those with a posterior inclusion probability (PIP) of greater than 0.95.

### Enrichment analysis

GOmeth (missMethyl v. 1.40.3) was used to assess for enrichment (GO and KEGG) for the significant MAJA CpGs (N_CpGs_ = 16), with the EPIC array CpGs tested (N_CpGs_ = 752,722) used as background. PANTHER v.19.0^52^ was used to assess for enrichment of the significant MAJA proteins (N_proteins_ = 46) with the input proteins (N_proteins_ = 133) as reference.

### EWAS catalogue

The EWAS catalogue (download date = 14/07/2025) was searched to identify any replication of previous Pollution EWASs. Initially, the catalogue was searched for CpGs matching high-confidence CpGs (N = 16) from our results and filtered for studies using “whole blood”. This revealed EWAS catalogue results for 15 CpGs, from 178 studies and 112 traits. These results were further filtered for studies with a study sample greater than 1000 and results where the p-value was less than the epigenome-wide significance threshold of 3.6 x 10^-8^, reducing the results to 15 CpGs and 36 traits from 60 studies. We assessed these results for previously reported CpG – pollutant associations and other traits associated with the CpGs in our results.

### MAJA Pollution & Proteomics Analysis

15,352 individuals had complete protein and pollution data in Generation Scotland. As for the EWAS, data were filtered to individuals with BMI >=16 and <=50 (N_remaining_ = 15,314). MAJA^103^ was again used to identify unique and shared associations between proteins, eight pollutants and smoking. Data preparation for the smoking pack-years was as above (N_missing_ = 361). Protein data were pre-regressed for covariates (age, sex, BMI, alcohol units per week and SIMD) using the same approach as for the methylation data (limma package (version 3.60.4))^78^ in R. Prior to regression, the same data transformations were applied and covariates were scaled (mean of 0, S.D. of 1) and any missing data (N_missing_ BMI = 135, SIMD rank = 484, alcohol units = 1427) were imputed using kNN (VIM v. 6.6.2)^102^. Residuals from the output of a linear model for each protein were scaled to have a mean of zero and unit variance prior to inclusion in the model. Prior distributions of effect sizes are assumed to have a multivariate spike and slab distribution to allow for markers with zero effect. 2000 model iterations were run, with 750 ‘burn-in’ iterations discarded prior to averaging effect sizes over the last 1250 posterior samples. Significant associations were taken to be those with a posterior inclusion probability (PIP) of greater than 0.95 and with a credible interval which does not cross 0.

As for the EWAS, comparative univariate analyses were also run for the proteomic data using GMRM-omi, the same parameters (prior mixture variance proportions set to 0.0, 0.001, 0.01, and 0.1, 2000 total iterations, 750 burn-in iterations, and exposures of 1-year, 6 months and 7 years).

### Identification of previous pollution-protein associations

A recent study in UK biobank investigated 2265 proteins associated with exposure to PM_2.5_ and NO_2_ in 51,357 participants^23^. We used this study as a comparator analysis. Using R to compare UniProt protein accessions, we identify the number of Generation Scotland mass-spectrometry proteins present in the UK BioBank Olink Explore platform and subsequently identify assess whether any of their identified significant associations are repeated in our analysis.

### Pollution EpiScore

Given that air pollution exposure can be both challenging to measure and computationally intensive to estimate, we sought to develop a biomarker of exposures using DNA methylation data from Generation Scotland. Participants who also took part in the STRADL (Stratifying Resilience and Depression Longitudinally) sub-cohort were reserved as a test subset (N_samples_ = 1,188)^67^. Blood samples for the STRADL sub-cohort were taken approximately 5 years after the GS baseline sampling. Baseline samples for those individuals and anyone from the same family pedigree were removed from the training subset, resulting in a training dataset number of 15,516^108^. We used the same multivariate approach as for the EWAS to generate a pollution EpiScore for the 8 modelled EMEP4UK pollutants (PM_2.5_, PM_10_, NO_2_, NO, NO_3_F_, NO_3_C_, SO_2_, O_3_). Methylation data were pre-regressed for age, sex and measurement batch, and the residuals of these linear models scaled and used for downstream analysis. Smoking pack-years was included as an outcome variable as previous; this variable was log-transformed (with +1 added to account for those with 0 smoking pack-years) and missing data (N = 331) imputed using kNN from the VIM package (v.6.2.2) in R. Pollutant data were scaled. The mean posterior CpG weights, calculated from post-burn-in iterations were extracted from the multivariate model.

We generated pollution EpiScores in two test sets, the STRADL cohort test-set (N unrelated and with methylation data = 695, Illumina EPIC v.1. array) and in LBC1936 participants (N = 895, Illumina 450k array). The CpG weights were projected onto measured DNA methylation in each test cohort to generate pollutant EpiScore values (the additive sum of all CpG weights multiplied by the measured CpG M-values). Pollution exposure estimates were generated for each test set using the same method as for the baseline GS participants. To assess whether these pollution EpiScores demonstrated an association with estimated air pollution exposure we correlated the EpiScores and exposure estimates for each test set and the eight pollutants. A cut-off of Pearson R > 0.1 and with P < 0.05 was used to select pollutant EpiScores which demonstrate an association with paired, estimated pollutant exposure.

## Data Availability

According to the terms of consent for Generation Scotland participants, access to data must be reviewed by the Generation Scotland Access Committee. Information on how to make an application can be found on the Generation Scotland website: https://genscot.ed.ac.uk/for-researchers/access. Lothian Birth Cohort data are available upon request from the LBC Study, University of Edinburgh (https://www.ed.ac.uk/lothian-birth-cohorts/data-access-collaboration).
Summary statistics generated during the study will be made available in Zenodo on publication. Code is available at https://github.com/marioni-group/GS_Molecular_markers_air_pollution

## Acknowledgements

Generation Scotland received core support from the Chief Scientist Office of the Scottish Government Health Directorates [CZD/16/6] and the Scottish Funding Council [HR03006] and is currently supported by the Wellcome Trust [216767/Z/19/Z]. Genotyping of the Generation Scotland samples was carried out by the Genetics Core Laboratory at the Edinburgh Clinical Research Facility, University of Edinburgh, Scotland and was funded by the Medical Research Council UK and the Wellcome Trust (Wellcome Trust Strategic Award “STratifying Resilience and Depression Longitudinally” (STRADL) Reference 104036/Z/14/Z). The DNA methylation profiling and analysis was supported by Wellcome Investigator Award 220857/Z/20/Z and Grant 104036/Z/14/Z (PI: Prof AM McIntosh) and through funding from NARSAD (Ref: 27404; awardee: Dr DM Howard) and the Royal College of Physicians of Edinburgh (Sim Fellowship; Awardee: Prof HC Whalley).

## Author contributions

JAR, SRC, TCR, JP and REM conceived and designed the study. JAR conducted all analyses. IK, JB, MR developed the two Bayesian ‘omics analysis methods. AR, DLM, SV, JC, SJT, MC, ADC, AG, HMS, CB, CBM, AZ and MR contributed to data generation. JAR drafted the initial manuscript. All authors read and approved the final manuscript.

## Funding

JAR is a University of Edinburgh Clinical Academic Track PhD student, supported by the Wellcome Trust (319878/Z/24/Z). HMS is a student on the University of Edinburgh Translational Neuroscience PhD programme funded by the Wellcome Trust (218493/Z/19/Z). ADC was supported by a Medical Research Council PhD Studentship in Precision Medicine with funding from the Medical Research Council Doctoral Training Program and the University of Edinburgh College of Medicine and Veterinary Medicine. REM and SRC are currently supported by BBSRC grant UKRI1940. For the purpose of open access, the author has applied a CC BY public copyright licence to any Author Accepted Manuscript version arising from this submission.

## Competing interests

C.B., A.Z. and M.R. are co-founders of Eliptica Ltd. C.B.M. is a consultant and shareholder of Eliptica Ltd (London, UK). R.E.M. is a scientific advisor to the Epigenetic Clock Development Foundation and Optima Partners. D.L.M. is employed by Optima Partners Ltd. The other authors have no competing interests to declare.

## Data availability

According to the terms of consent for Generation Scotland participants, access to data must be reviewed by the Generation Scotland Access Committee. Information on how to make an application can be found on the Generation Scotland website: https://genscot.ed.ac.uk/for-researchers/access. Lothian Birth Cohort data are available upon request from the LBC Study, University of Edinburgh (https://www.ed.ac.uk/lothian-birth-cohorts/data-access-collaboration). Summary statistics generated during the study are available in Zenodo under a Creative Commons Attribution 4.0 International license.

## Code availability

All code associated with this manuscript is available open access on GitHub under the GNU General Public License version 3.0 () and Zenodo

## Notes

### Author Declarations

The NHS Tayside Committee on Medical Research Ethics (REC Reference Number: 05/S1401/89) gave ethical approval for the Generation Scotland cohort and Research Tissue Bank status was granted by the East of Scotland Research Ethics Service (REC Reference Number: 20/ES/0021). The Lothian Research Ethics committee (LREC/2003/2/29) and Multi-Centre Research Ethics Committee for Scotland (MREC/01/0/56) gave ethical approval for the LBC1936 study.

