## Supplementary Methods for "Air pollution exposure in Generation Scotland: molecular fingerprints and health outcomes"

**EWAS air pollution: Supplementary Methods**

1. **DNA methylation quality control (Generation Scotland)**

DNA methylation was profiled in four waves (2016-2021). The quality control steps have previously been described in full^1^. Briefly, the steps were as follows: visual inspection of log median intensity of methylated versus unmethylated signal for each array was used to identify and remove technical outliers. Samples were removed where methylation-derived sex (ShinyMethyl vs.1.10.0 (wave 1), 1.14.0 and 1.30.0 (waves 2-4)^2^) disagreed with self-reported sex. In wave 1, samples were removed where >1% of CpG sites had a detection p-value of > 0.05 (pfilter function, watermelon v.1.18.0^3^) and probes were removed if the beadcount was <3 in >5% of samples, or if >0.5% of the samples had a detection P-value of > 0.05 (shinyMethyl v1.10.0). In waves 2-4, samples were removed where the median methylated signal intensity was > 3 standard deviations lower than expected, and if > 0.5% CpGs in the sample had a detection p-value > 0.01. Poorly performing probes were removed if the bead-count was <3 in >5% of samples, or if >1% had a detection p-value of >0.01 (Meffil vs. 1.10 and 1.12^4^).

2. **Cox models data preparation health outcome definition**

Data preparation for the Cox mixed-effects models is detailed in the table below (**Table 1)** for each considered disease. Incident and prevalent disease were defined using ICD codes in electronic health-care records from secondary care. Dementia diagnoses were determined through a combination of GP and secondary care data (7,580 of 21,725 participants had GP data available which translated to 3,714 of 10,761 after age-filtering and removal of prevalent cases, see below). Demographic summaries can be found in **supplementary table 25** and disease definitions (with ICD codes) in **supplementary table 28**. For each condition, the starting sample size was 22,071. For cardiovascular disease (CVD), ischaemic heart disease (IHD), myocardial infarction (MI) and ischaemic stroke (stroke), data were filtered to ages between 40 and 69, in alignment with the SCORE2 clinical risk predictor^5^. For dementia, data were filtered to ages greater than or equal to 65 at diagnosis or at censor to remove any anomalous cases such as early-onset. The censor dates were set to October 2024 (dementia diagnosis) or October 2023 (CVD, IHD, MI, stroke, hypertension) as the last dates for cross-referencing with the relevant health records. The first date of the relevant diagnosis is recorded as disease onset. Missing variables were imputed using kNN (VIM v.6.6.2, k = 5). Covariates are defined as follows: BMI = body mass index (kg/m^2^); smoking = smoking pack-years (one smoking pack-year equates to smoking 20 cigarettes per day per year); alcohol = alcohol units consumed per week, SIMD = Scottish Index of Multiple Deprivation (rank, 2009)^6^; HDL = high density lipoproteins (mmol/L); total cholesterol (mmol/L).

On initial exploration of the data, assumptions of proportional hazards (cox.zph function from the survival package v.3.8.3^7^) were violated for 4 of the 8 pollutants (CVD) and 3 of 8 pollutants (Dementia). We therefore explored hazard ratios at different follow-up times and with the pollutant as a categorical variable, divided into “high” (> mean) vs “low” (< mean). Using a categorical pollutant variable largely resolved failures of the proportional hazards assumption and we therefore use this for the remainder of the disease-association analysis (See **Supplementary figures 11 and 12**). For the dementia models, removing APOEe4 allele count as a covariate resolved violation of proportional hazards with minimal impact on the estimated hazard ratio and confidence interval (**Supplementary figure 12).**

|  | **CVD** | **IHD** | **MI** | **Stroke** | **Hypertension** | **Dementia** |
| --- | --- | --- | --- | --- | --- | --- |
| **Data Filtering (N remaining)** | Prevalent CVD removed (21,388)  BMI ≥ 16 and ≤ 50  (21,332)  Age ≥40 and ≤69 (13,023) | As for CVD | As for CVD | As for CVD | Prevalent CVD removed (21,388)  BMI ≥ 16 and ≤ 50 (21,332)  Prevalent HTN removed (20,753) | Age ≥ 65 (10,770)  Remove prevalent cases (10,761)  BMI ≥ 16 and ≤ 50 (10, 739) |
| **N missing covariates**  **(imputed via kNN)** | BMI: 647  Smoking: 313  Alcohol: 1210  SIMD rank: 231  HDL: 1114  Total cholesterol: 1095 | As for CVD | As for CVD | As for CVD | BMI: 1270  Smoking: 569  Alcohol: 1963  SIMD rank: 664  HDL: 2202  Total cholesterol: 2164 | BMI: 736  Smoking: 243  Alcohol: 1134  SIMD rank: 154 |
| **Final N** | 13,023 | 13,023 | 13,023 | 13,023 | 20,753 | 10,739 |
| **N Cases** | 905 | 530 | 145 | 159 | 1689 | 323 |
| **Covariates (full model)** | Age, sex, kinship, BMI, alcohol consumption, SIMD rank, HDL, total cholesterol, prevalent diabetes, prevalent hypertension | As for CVD | As for CVD | As for CVD | Age, sex, kinship, BMI, alcohol consumption, SIMD rank, HDL, total cholesterol, prevalent diabetes | Age, sex, kinship, BMI, alcohol consumption, SIMD rank, **APOEe4 allele count**, HDL, total cholesterol, prevalent diabetes, prevalent hypertension |

**Table. 1. Data preparation for mixed-effects Cox models.**

6.	Scottish Government. Scottish Index of Multiple Deprivation 2009, General Report.

7.	Therneau, T. *A Package for Survival Analysis in R. R Package Version 3.5-8*. (2024).
