## Supplementary Figures for "Air pollution exposure in Generation Scotland: molecular fingerprints and health outcomes"

**EWAS air pollution – Supplementary figures**

**Figure 1. Pollution concentrations across exposure over the GS recruitment period**

**A**. Average annual pollutant levels across Scotland. **B**. Number of daily exceedances of WHO limits (2021) for NO_2_, PM_10_, PM_2.5_, SO_2_ across Scotland.

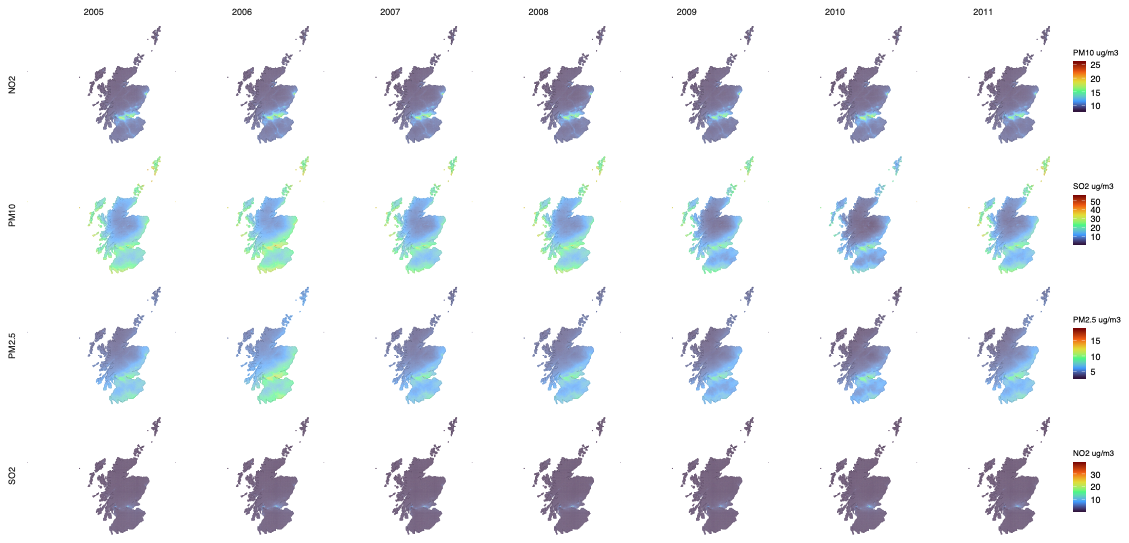

A

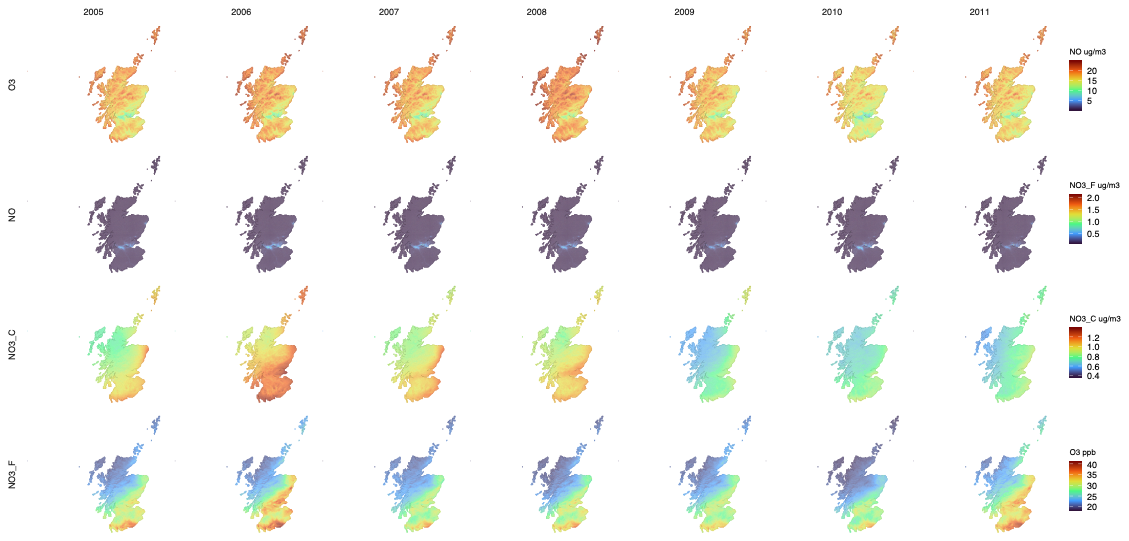

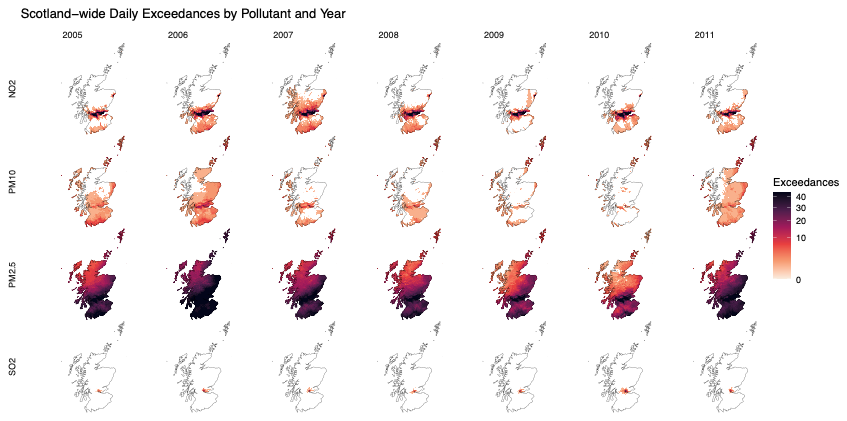

B

**Figure. 2. Spatial correlations of unassigned EMEP4UK data.**

Analysis carried out using (Terra package in R v. 1.8.29). EMEP4UK data was cropped to Scotland and yearly averages for each cell in the raster calculated. Correlations between years were then assessed using the layerCor function. ComplexHeatmap (v. 2.16.0) was then used to generate heatmaps of the yearly spatial correlations for each pollutant.

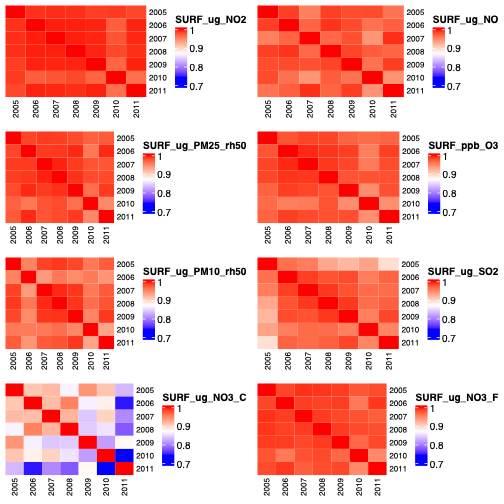

**Figure 3. Comparison of exposure of different averaging time frames (GS)**

Average pollution exposure for each pollutant by different averaging time frames. 6 month refers to 183 days prior to the baseline appointment, 1 year refers to 365 days prior to the baseline appointment and 7 years refers to an average over the recruitment period for Generation Scotland and one-year prior to this (2005 – 2011).

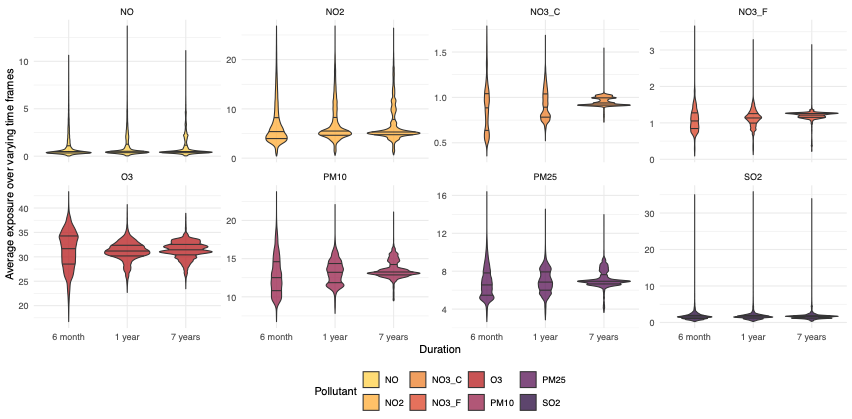

**Fig. 4. Between pollutant correlations for Pollutant exposure**

Input data comprised of 365-day exposure pre-baseline appointmen*t.* Created using cor function from ComplexHeatmap (v. 2.16.0) in R.

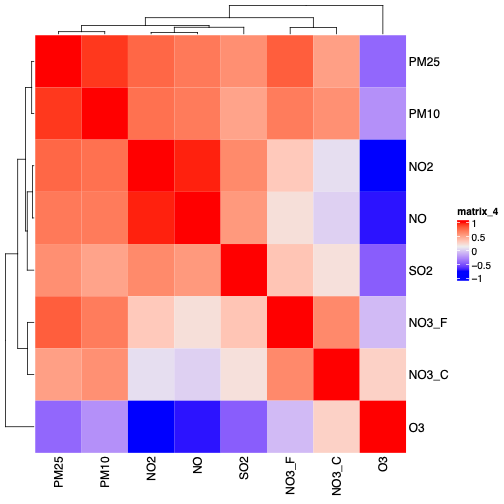

**Figure 5. Distribution of number of 24hr exceedances of WHO limits for 365-day pollution exposure.** The World Health Organisation (WHO, 2021) set 24hr limits for ambient air pollutant concentrations of four main pollutants: NO_2_, PM_10_, PM_2.5_ and SO_2_. This plot demonstrates number of exceedance days for GS participants. Counts of participants with greater than 4 exceedance days are coloured in darker blue as the 99^th^ percentile of daily values exceeding the limit.

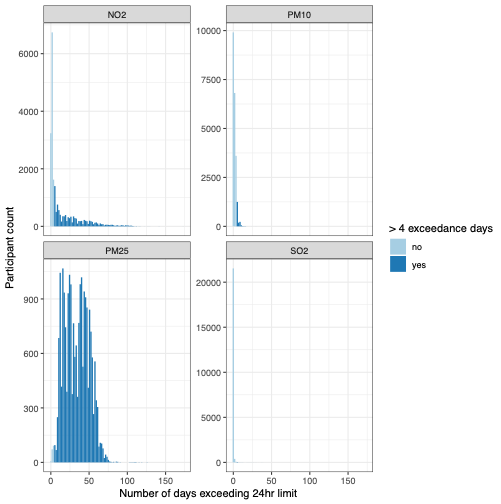

**Figure 6. Comparison of one-year exposures between GS, LBC and STRADL**

One-year (365-day) average pre-baseline sample exposures were calculated for Generation Scotland (GS), Lothian Birth Cohort 1936 (LBC) and Stratifying Resilience and Depression Longitudinally (STRADL) participants. Violin plots demonstrate the spread of these averages between cohorts. Interquartile ranges and medians are denoted by horizontal lines within each violin plot.

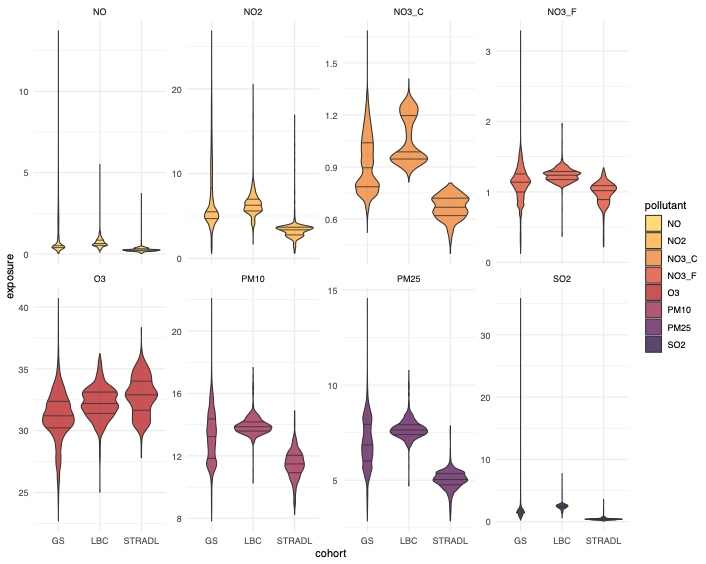

**Figure. 7. Principal components analysis of pollution data.**

Principal components of the assigned pollutant exposure (N = 22,071) were calculated using the PCAtools package (v. 2.12.0), in R. (See also **Supplementary Table x.**).

A. Scree plot of pollution data, demonstrating that the first two principal components constitute around 80% of the variance.
B. Biplot of PC analysis.

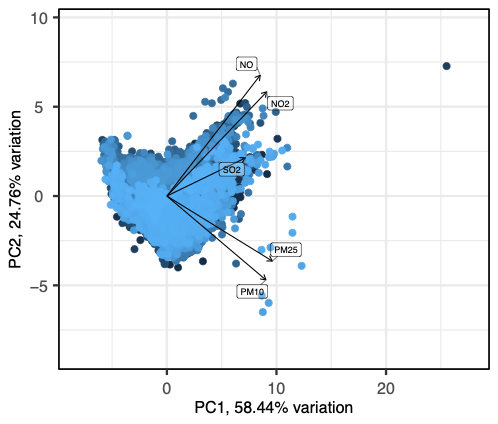

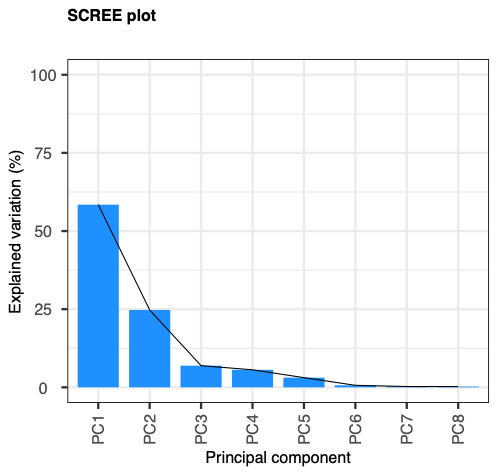

**Figure 8. Cox model results of pollutant principal components and health outcomes**

The first two principal components (accounting for >80% of the variance in the pollutant data) were assessed in association with incident disease. P_Bonferroni_ was set at 0.05/(2*6), to account for the number of principal components and diseases compared.

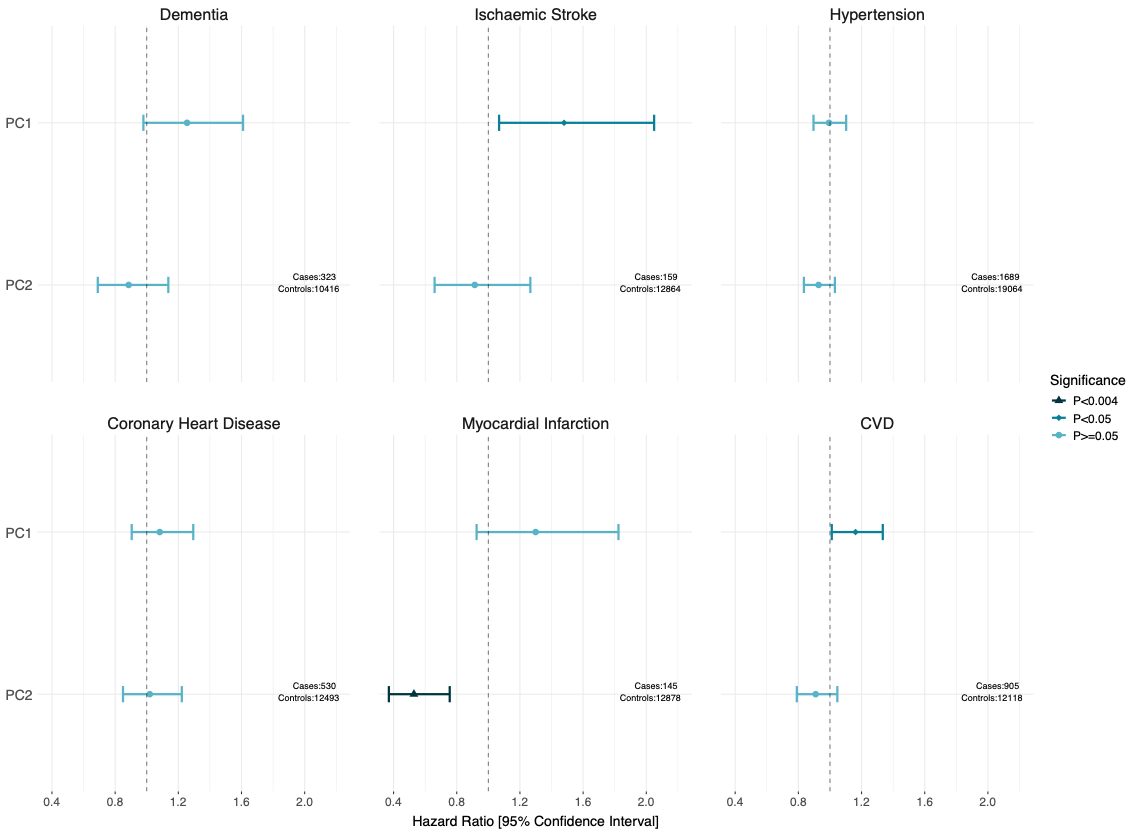

**Figure 9. Results from Age-acceleration ~ pollutant associations**

**A.** Comparing associations across different pollutant averaging times. Fully adjusted model used. Pollutant ~ Age acceleration + age + sex + kinship + log(alcohol units) + log(bmi) + simdrank + log(smoking pack_years) + white cell proportions. **B** Results from Age-acceleration ~ pollutant associations for all models and one-year exposure data.

Model 1 = Pollutant ~ Age acceleration + age + sex + kinship; Model 2 = Pollutant ~ Age acceleration + age + sex + kinship + log(alcohol units) + log(bmi) + simdrank; Model 3 = Pollutant ~ Age acceleration + age + sex + kinship + log(alcohol units) + log(bmi) + simdrank + log(smoking pack_years); Model 4 = Pollutant ~ Age acceleration + age + sex + kinship + log(alcohol units) + log(bmi) + simdrank + log(smoking pack_years) + white cell proportions.

A

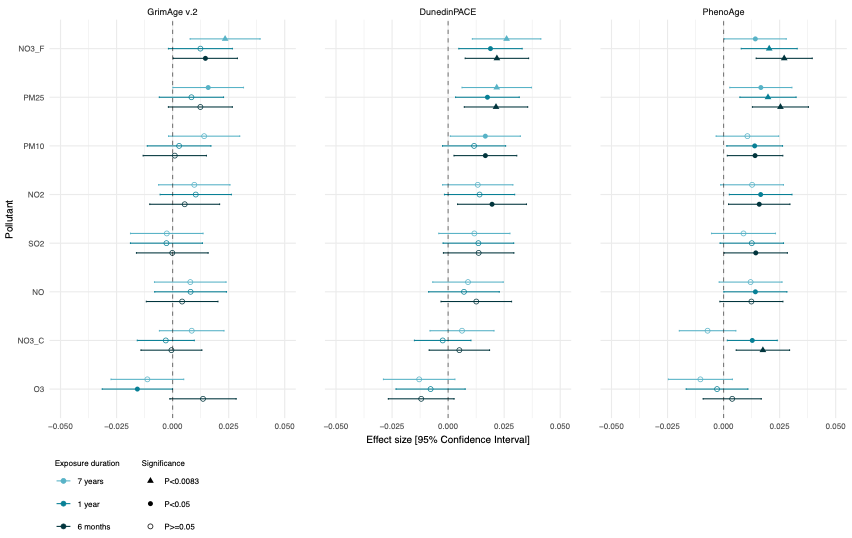

B

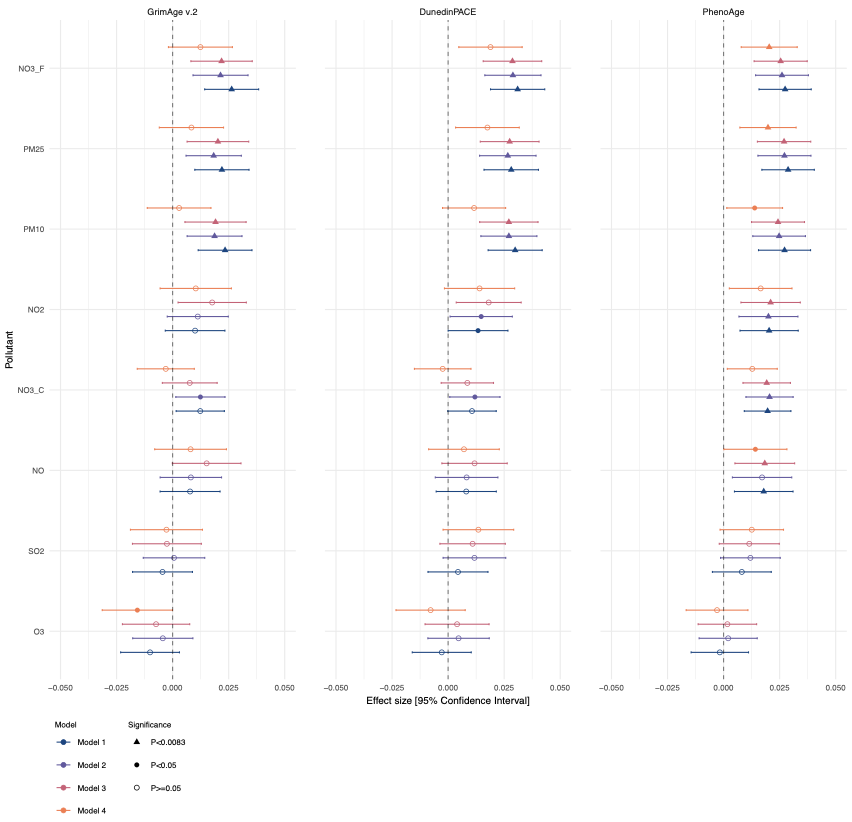

**Fig 10. String DB network of 12 unique genes from pollution-associated CpGs.**

Creation & download date: 12/09/25. Legend is a screenshot of the StringDB legend from the same date.

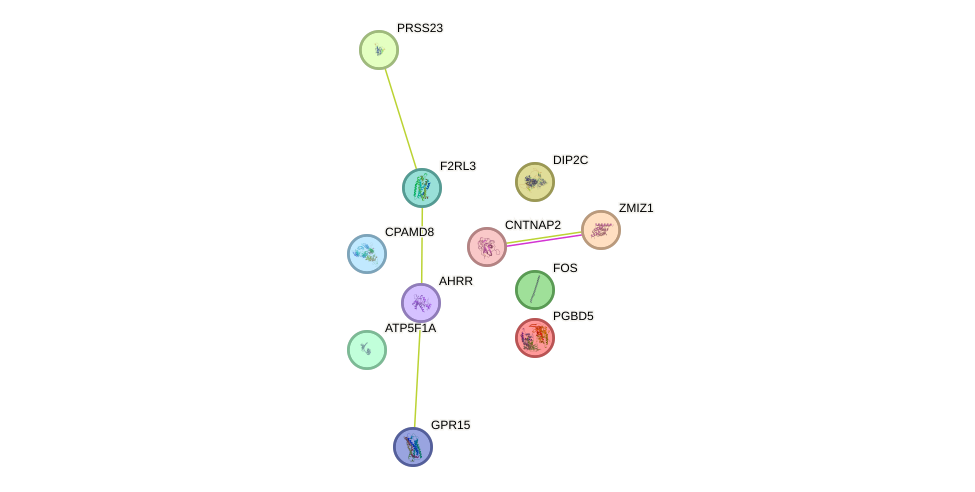

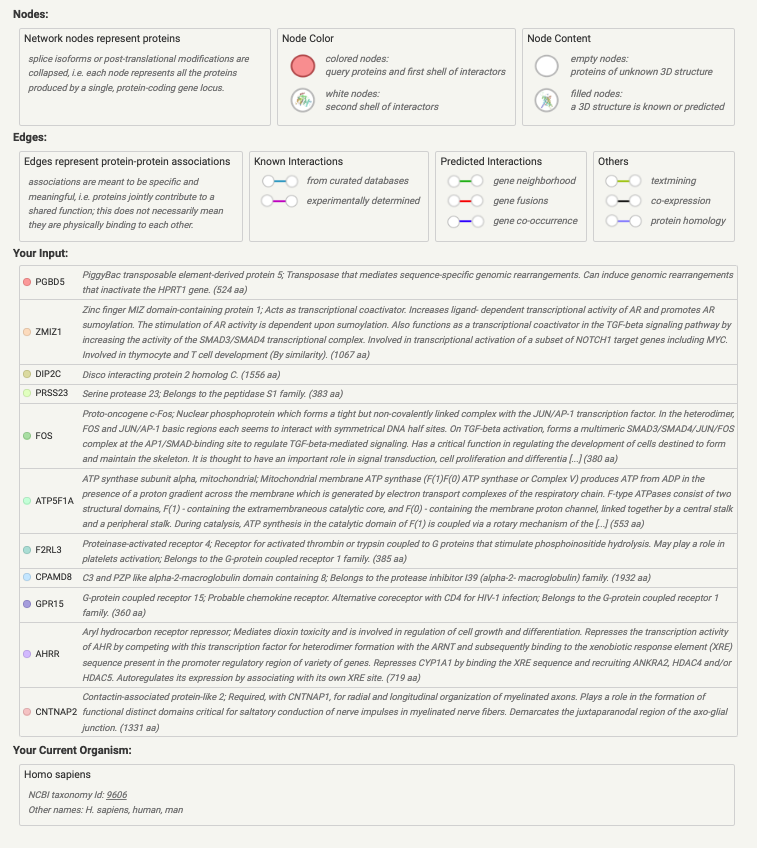

**Figure 11. Comparison of results between multivariate (MAJA) and univariate (GMRM) EWAS.** A. Comparison of effect sizes for all associations found with multivariate and univariate approaches. B. Variance explained and number of CpG associations with PIP > 0.95 for each method and duration of exposure. C. Correlation of effect sizes between multivariate and univariate approaches for 1-yr data.

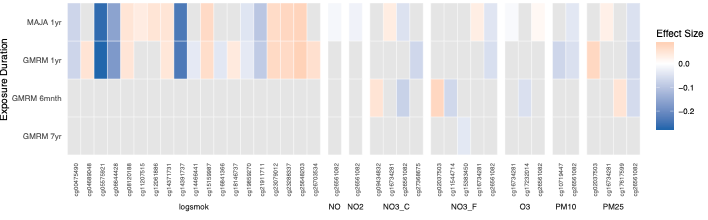

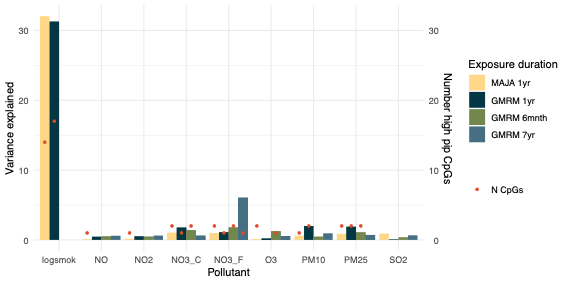

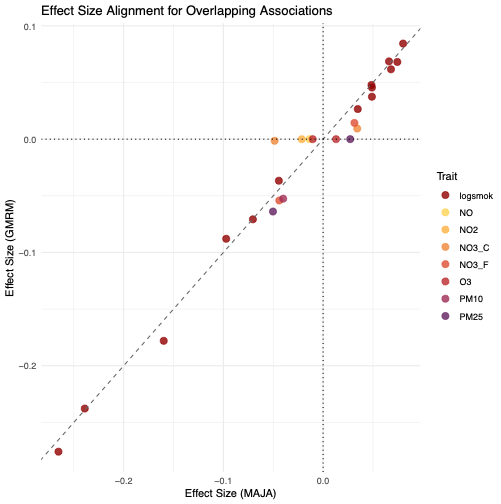

A

B

C

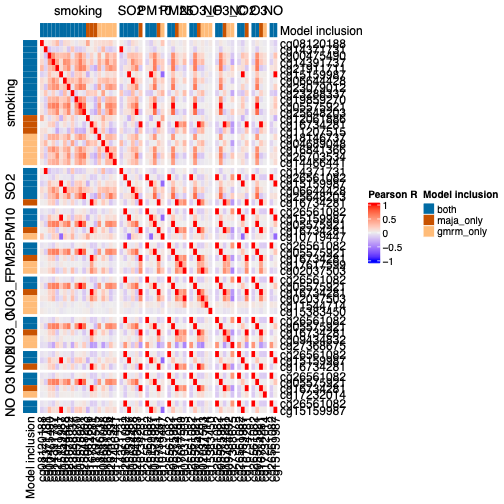

**Figure 12. Comparison of results between multivariate (MAJA) and univariate (GMRM) PWAS.** A. Correlation of effect sizes between multivariate and univariate approaches for 1-yr data. B. Variance explained and number of CpG associations with PIP > 0.95 for each method and duration of exposure. C. Comparison of effect sizes for all associations found with multivariate and univariate approaches.

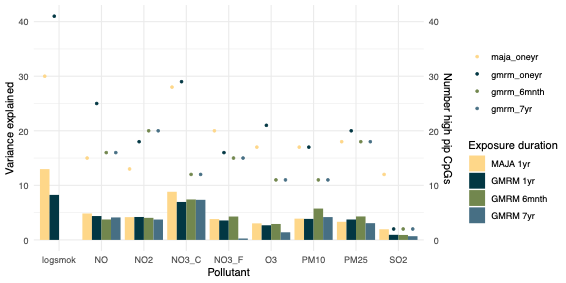

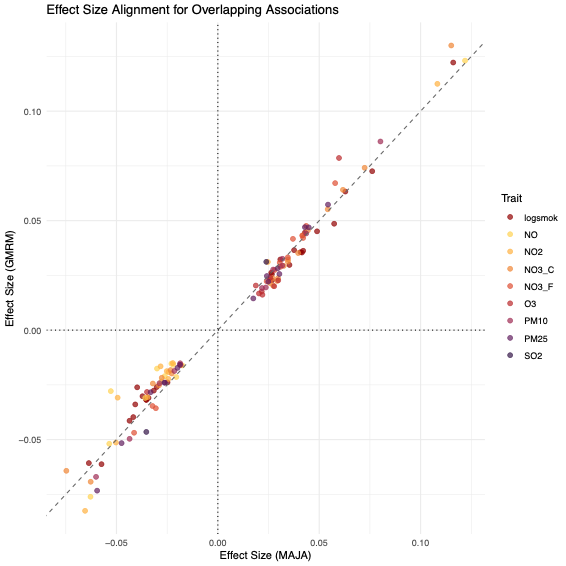

A

B

C

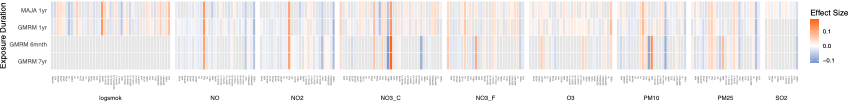

**Figure 13. Coxph models of incident CVD and pollutant exposure.** A. Pollutant as a continuous variable, with exploration of proportional hazards across different follow-up periods**.** B. Pollutant as a categorical variable (high vs. low), with exploration of proportional hazards across different follow-up periods

A

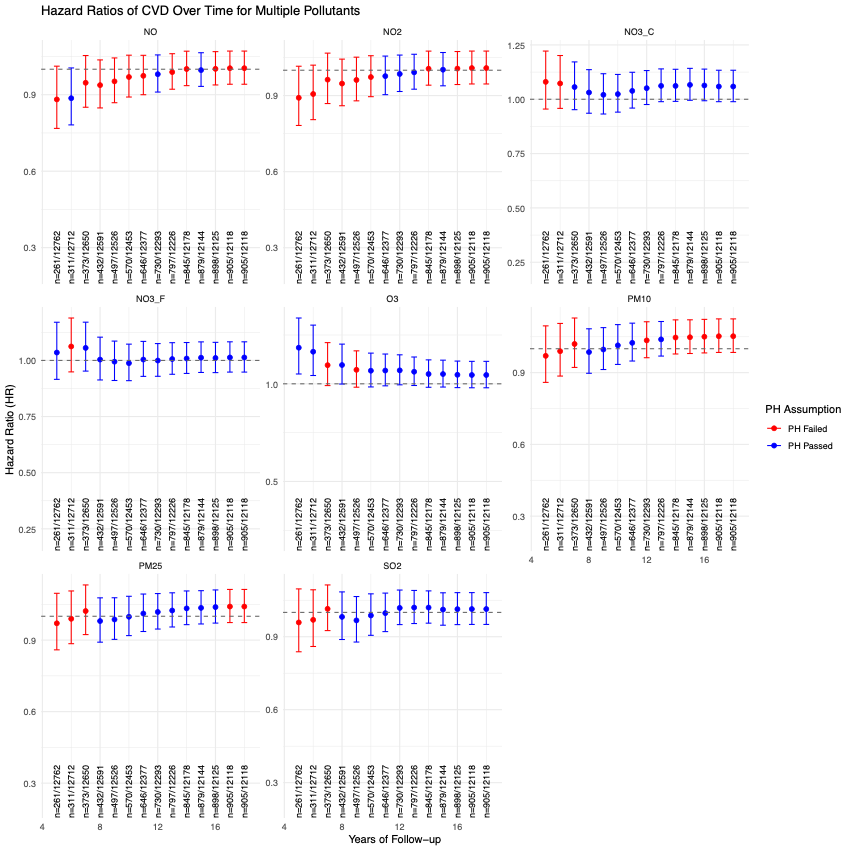

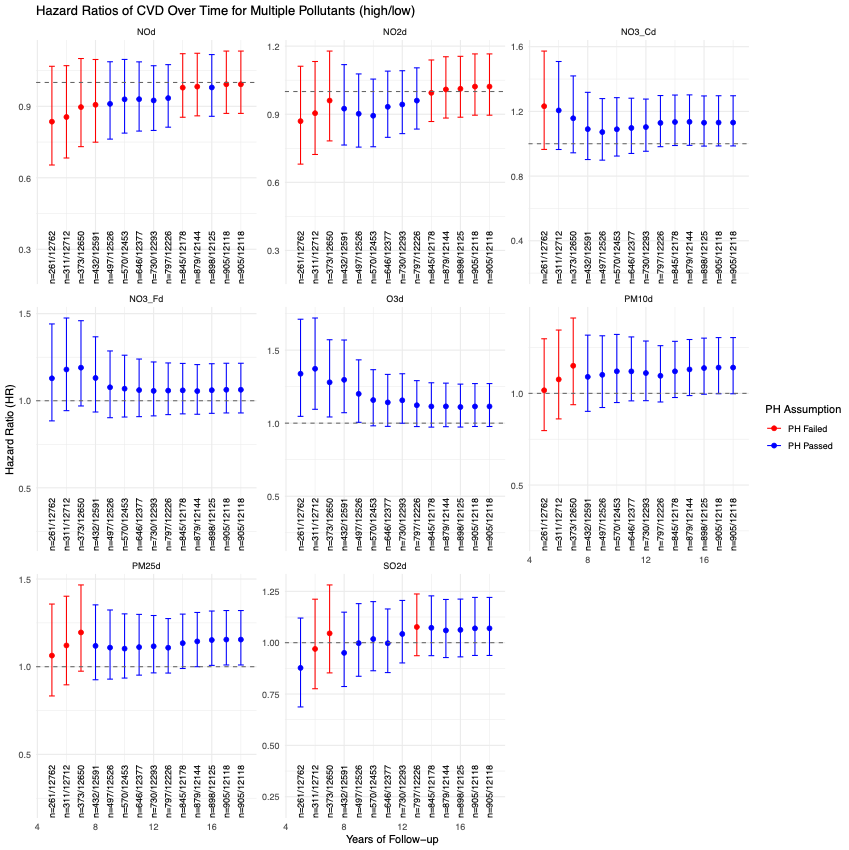

B

**Fig 14. Coxph models of incident dementia and pollution exposure.** A. Pollutant as a continuous variable, with exploration of proportional hazards (using coxph models) across different follow-up periods for the fully adjusted model. B. Pollutant as a categorical variable (high vs. low), with exploration of proportional hazards (using coxph models) across different follow-up periods for fully adjusted model. C. Pollutant as a categorical variable (high vs. low), with exploration of proportional hazards (using coxph models) across different follow-up periods for fully adjusted model, excluding APOEe4 allele count as a covariate. Removing this covariate mostly resolves proportional hazards but minimally effects the hazard ratio estimates.

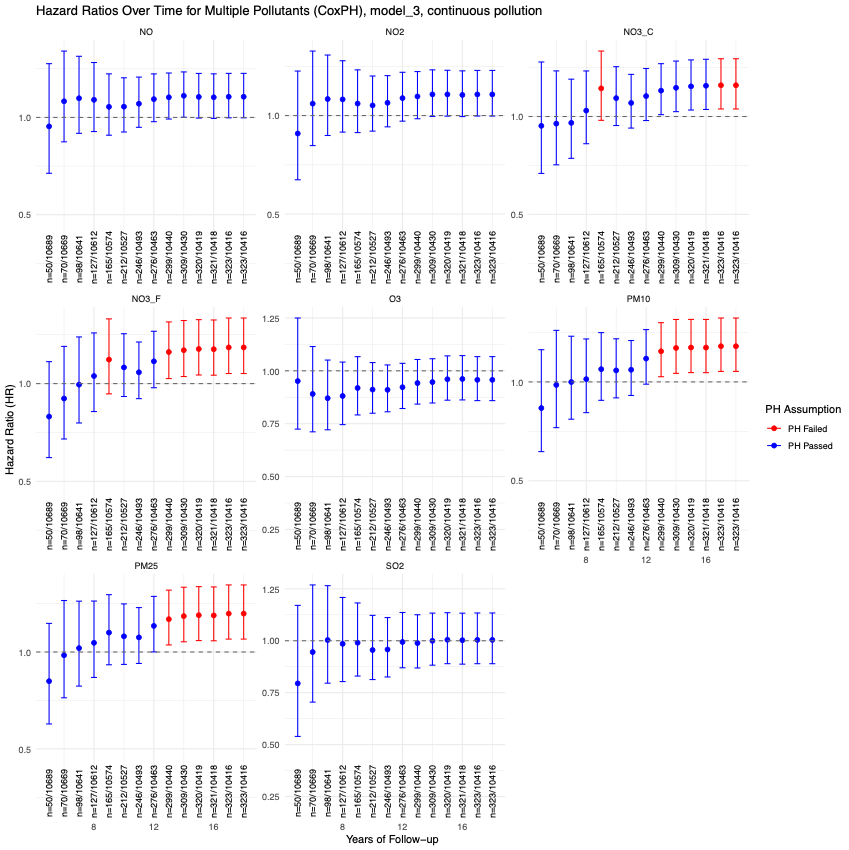

A

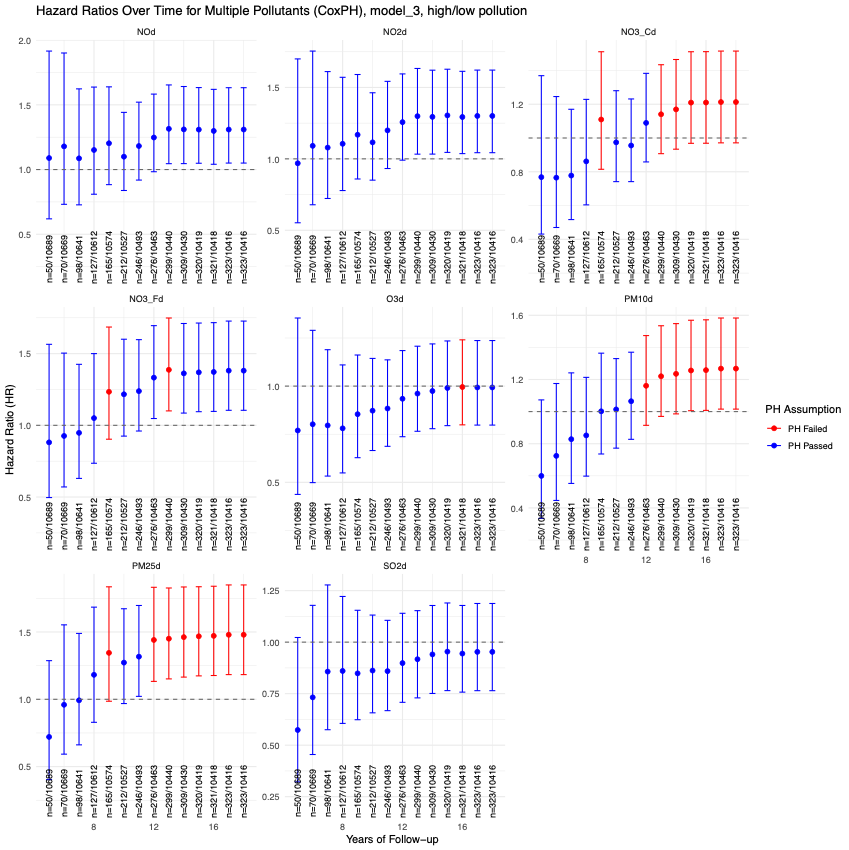

B

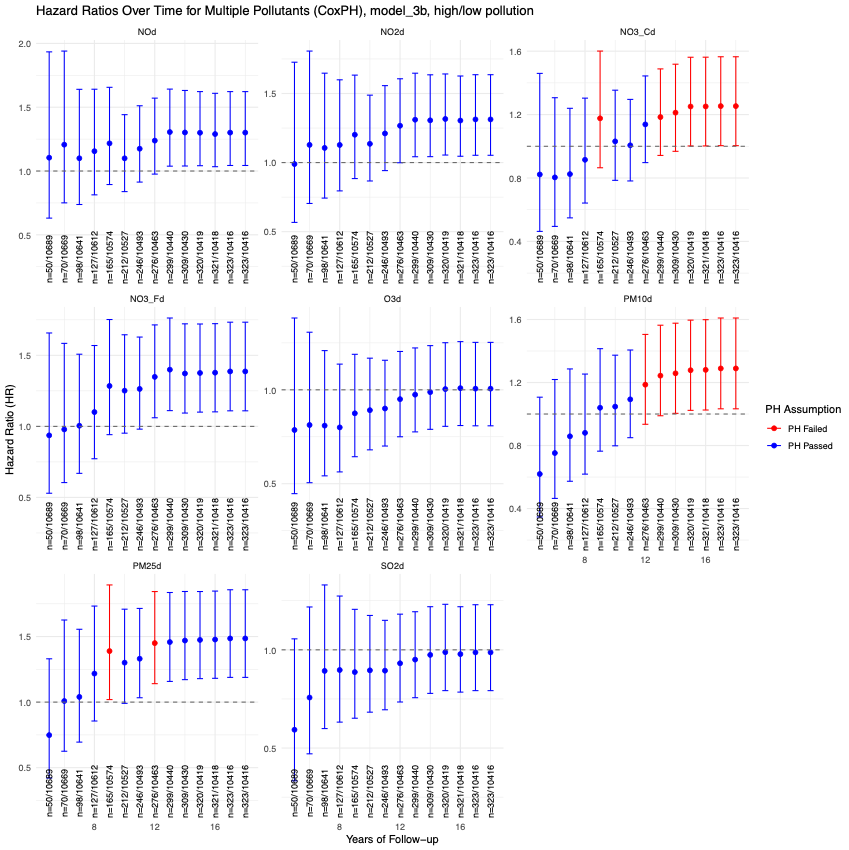

C

**Fig. 15 Correlation between pollutants and covariates**

Correlation between pollutants and included covariates for 18,556 participants (N for Epi-Age analysis). Covariates were imputed where data were missing (see methods). Heatmap constructed using ComplexHeatmap in R.

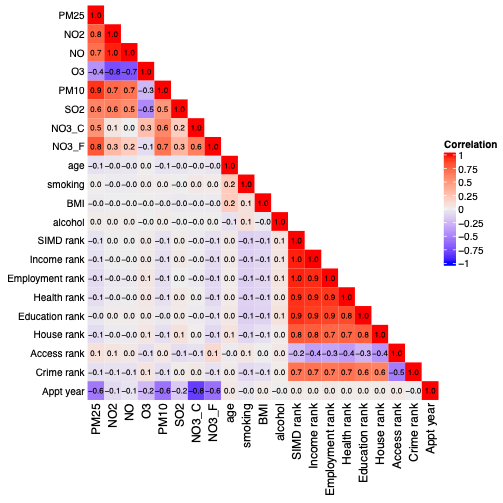
